# Multi-model ensembles in infectious disease and public health: Methods, interpretation, and implementation in R

**DOI:** 10.1101/2024.06.24.24309416

**Authors:** Li Shandross, Emily Howerton, Lucie Contamin, Harry Hochheiser, Anna Krystalli, Consortium of Infectious Disease Modeling Hubs, Nicholas G. Reich, Evan L. Ray

## Abstract

Combining predictions from multiple models into an ensemble is a widely used practice across many fields with demonstrated performance benefits. Popularized through domains such as weather forecasting and climate modeling, multi-model ensembles are becoming increasingly common in public health and biological applications. For example, multi-model outbreak forecasting provides more accurate and reliable information about the timing and burden of infectious disease outbreaks to public health officials and medical practitioners. Yet, understanding and interpreting multi-model ensemble results can be difficult, as there are a diversity of methods proposed in the literature with no clear consensus on which is best. Moreover, a lack of standard, easy-to-use software implementations impedes the generation of multi-model ensembles in practice. To address these challenges, we provide an introduction to the statistical foundations of applied probabilistic forecasting, including the role of multi-model ensembles. We introduce the `hubEnsembles` package, a flexible framework for ensembling various types of predictions using a range of methods. Finally, we present a tutorial and case-study of ensemble methods using the `hubEnsembles` package on a subset of real, publicly available data from the FluSight Forecast Hub.

## 1 Introduction

Predictions of future outcomes are essential to planning and decision making, yet generating reliable predictions of the future is challenging. One method for overcoming this challenge is combining predictions across multiple, independent models. These combination methods (also called aggregation or ensembling) have been repeatedly shown to produce predictions that are more accurate^1,2^ and more consistent^3^ than individual models. Because of the clear performance benefits, multi-model ensembles are a widely used statistical tool across fields, including weather forecasting^4^, climate modeling^5^, and economics^6^. In the last decade, the number of multi-model ensemble predictions generated and used in real time for public health planning and response has grown rapidly.

In particular, predicting infectious disease outbreaks and anticipating the effects of potential interventions has demonstrated utility for public health officials and medical practitioners. Underlying these predictions are mathematical models that use historical disease incidence data to make probabilistic predictions of incidence in the future^7–11^. Given the performance benefits of multi-model ensembles, it is becoming increasingly common to convene multiple modeling teams into a collaborative “hub”^12^, where each team generates independent predictions that are aggregated to collectively produce an ensemble. For example, this approach has been used to make real-time, multi-model predictions for seasonal influenza^10,13^, dengue^14^, West Nile virus^15^, and more recently SARS-CoV-2^16–18^.

Generating multi-model ensembles or interpreting the resulting predictions depends on understanding the underlying statistical methodology. There are many proposed methods for generating ensembles, and these methods differ in at least one of two ways: (1) the function used to combine or “average” predictions, and (2) how predictions are weighted when performing the combination. A few methodological papers have discussed theory of multi-model ensembles and tested various ensembling methods in public health applications specifically^19–25^, yet there is no consensus on which method should be favored. There are software packages that support various aspects of multi-model ensembling^26–29^. However, these packages only support a subset of methods and prediction types, illustrating the need for standard, easy-to-use implementations of common methods.

Here, we provide an introduction to the statistical foundations of multi-model ensembles in applied probabilistic forecasting (Section 2). In addition, to improve accessibility, reproducibility, and interoperability, we have developed a comprehensive R package `hubEnsembles` that implements these methods. The `hubEnsembles` package provides a flexible framework for generating ensemble predictions from multiple models across a range of common methods and prediction types; it is situated within the broader “hubverse” collection of open-source software and data tools to facilitate the development and management of collaborative modeling exercises^30^. These two factors together, a simple implementation framework across methods and integration with hubverse data standards and tools, make `hubEnsembles` accessible and easy to use. Finally, we present a basic demonstration of multi-model ensemble generation and interpretation (Section 4), and a more in-depth analysis using real influenza forecasts (Section 5). Together, these case studies motivate a discussion and comparison of the various methods (Section 6). While the case studies focus on infectious disease applications, the software and tools presented are general and could be used for applications in other areas of biomedical and public health research, or other domains. By reviewing multi-model ensemble methodology and synthesizing these methods into an easy-to use implementation, this tutorial provides guidance on understanding, interpreting, and implementing multi-model ensembles.

## 2 How to generate a multi-model ensemble

In this section, we provide an overview of the process to generate a multi-model ensemble, including basic definitions of key statistical concepts in probabilistic forecasting, and an overview of the classes of methods that are typically used for generating a multi-model ensemble.

### 2.1 Key statistical concepts in forecasting

Generating an ensemble requires multiple predictions to be combined, and a combination method for calculating the ensemble from these predictions (Figure 1). These predictions will often be produced by different statistical or mathematical models, and the output from these models (referred to as “model output” from here on) will vary based on the setting. For example, some public health questions, such as short-term resource allocation, may depend on a forecast of public health outcomes weeks into the future, whereas longer-term decisions about vaccination schedules may require projections months into the future across multiple possible scenarios. Some intervention decisions, such as quarantine and isolation policy, may depend on estimates of key biological parameters such as the generation interval for an infectious pathogen. Throughout, we will use the general term “prediction” to encapsulate all such outcomes that could be modeled. Predictions can also capture varying degrees of uncertainty in the outcome. A *point prediction* gives a single estimate of an outcome while a *probabilistic prediction* provides an estimated probability distribution over a set of outcomes. In either case, the basic steps required to generate an ensemble are the same.

**Figure 1:**
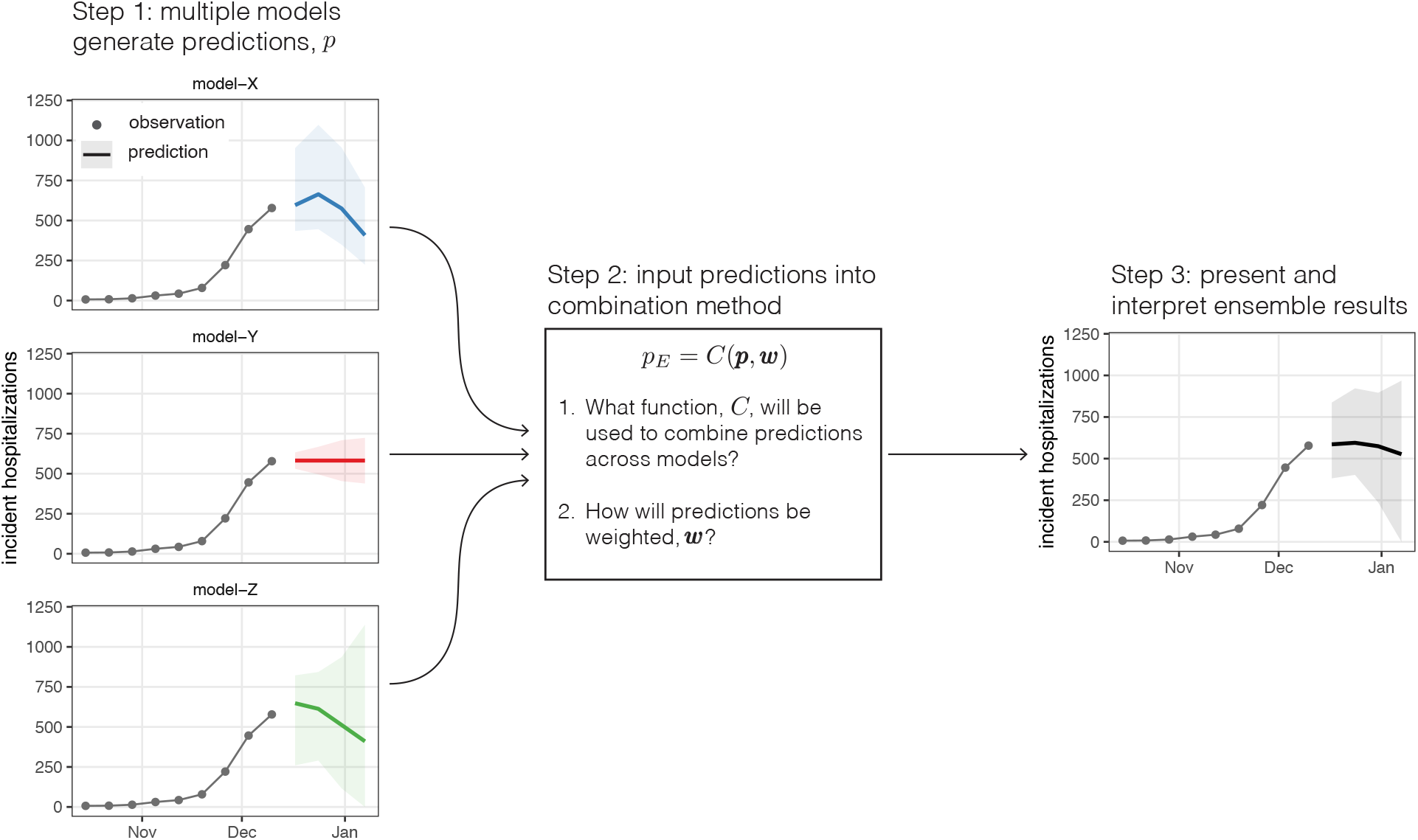
Overview of process to generate a multi-model ensemble. Predictions, *p*_*i*_ are generated from *N* independent models (step 1). Then those predictions, *p* = {*p*_*i*_|*i* ∈ 1, …, *N*}, are combined with some function, *C*, and set of weights, *w* = {*w*_*i*_|*i* ∈ 1, …, *N*}. This figure illustrates example probabilistic forecasts for incident influenza hospitalizations, where the median (line) and 90% prediction interval are shown. In this case, the ensemble is constructed using the linear pool method (*F*_*LOP*_ (*x*)}).

### 2.2 Mathematical definitions and properties of ensemble methods

Here, we use *N* to denote the total number of individual predictions that the ensemble will combine. For example, if predictions are produced by different models, *N* is the total number of models that have provided predictions. Individual predictions will be indexed by the subscript *i*. Optionally, one can calculate an ensemble that uses a weight *w*_*i*_ for each prediction; we define the set of model-specific weights as *w* = {*w*_*i*_|*i* ∈ 1, …, *N*}. Informally, predictions with a larger weight have a greater influence on the ensemble prediction, though the details of this depend on the ensemble method (described further below).

Then, for a set of *N* point predictions, *p* = {*p*_*i*_|*i* ∈ 1, …, *N*}, each from a distinct model *i*, an ensemble of these predictions is

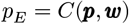

using any function *C* and any set of model-specific weights *w*. For example, an arithmetic average of predictions yields 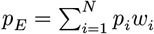, where the weights are non-negative and sum to 1. If *w*_*i*_ = 1/*N* for all *i*, all predictions will be equally weighted. More complex functions for aggregation are also possible, such as a (weighted) median or geometric mean.

For probabilistic predictions, there are two commonly used classes of methods to average or ensemble multiple predictions: quantile averaging (also called a Vincent average^31^) and probability averaging (also called a distributional mixture or linear opinion pool^32^)^33^. To define these two classes of methods, let *F* (*x*) be a cumulative density function (CDF) defined over values *x* of the target variable for the prediction, and *F* ^−1^(*θ*) be the corresponding quantile function defined over quantile levels *θ* ∈ [0, 1]. Throughout this article, we may refer to *x* as either a ‘value of the target variable’ or a ‘quantile’ depending on the context, and similarly we may refer to *θ* as either a ‘quantile level’ or a ‘(cumulative) probability’. Additionally, we will use *f*(*x*) to denote a probability mass function (PMF) for a prediction of a discrete variable or a discretization (such as binned values) of a continuous variable.

The quantile average combines a set of quantile functions,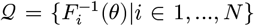, with a given set of weights, *w*, as

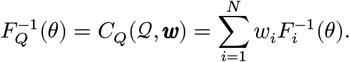

This computes the average value of predictions across different models for each fixed quantile level *θ*. For a normal distribution or any distribution with a location and scale parameter, the resulting quantile average will be the same type of distribution, with location and scale parameters that are the average of the location and scale parameters from the individual distributions (Figure 2, panel B). In other words, this method interprets the predictive probability distributions that are being combined as uncertain estimates of a single true distribution. It is also possible to use other combination functions, such as a weighted median, to combine quantile predictions.

**Figure 2:**
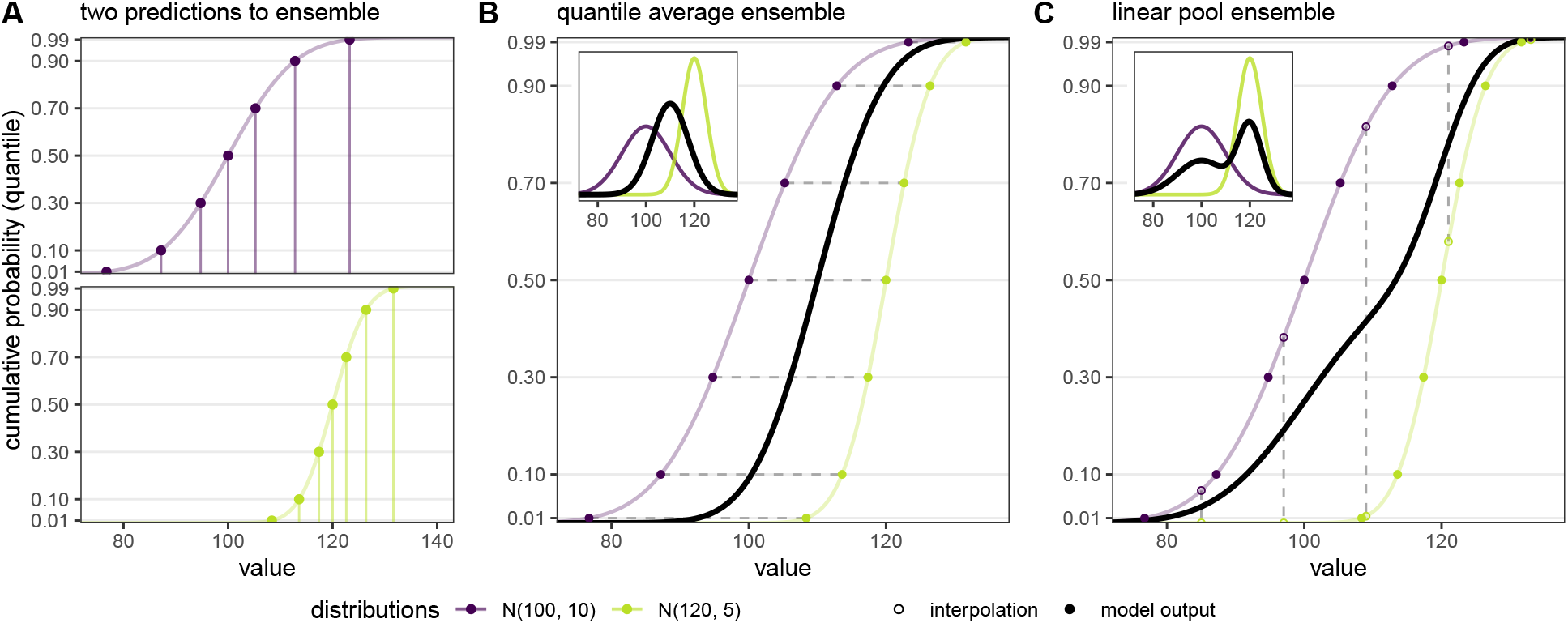
(Panel A) Example predictions from two distributions (*N*(100, 10) in purple and *N*(120, 5) in green) shown as cumulative distribution functions (CDFs). To ease submission to a hub, the prediction can be summarized at a fixed number of points along the distribution. Here, the solid points show model output for seven fixed quantile levels (*θ* = 0.01, 0.1, 0.3, 0.5, 0.7, 0.9, and 0.99). The y-axis ticks show each of the fixed quantile levels. The associated values for each fixed quantile level are shown with vertical lines. (Panel B) Quantile average ensemble, which is calculated by averaging values for each fixed quantile level (represented by horizontal dashed gray lines). The distributions and corresponding model outputs from panel A are re-plotted and the black line shows the resulting quantile average ensemble. Inset shows corresponding probability density functions (PDFs). (Panel C) Linear pool ensemble, which is calculated by averaging cumulative probabilities for each fixed value (represented by vertical dashed gray lines). The distributions and corresponding model outputs from panel A are re-plotted. To calculate the linear pool in this case, where model outputs are not defined for the same values (i.e., vertical lines in Panel A do not line up), the model outputs are used to interpolate the full CDF for each distribution from which predicted cumulative probabilities can be extracted for fixed values (shown with open circles). The black line shows the resulting linear pool average ensemble. Inset shows corresponding PDFs.

The probability average or linear pool is calculated by averaging probabilities across predictions for a fixed value of the target variable, *x*. In other words, for a set of CDFs *ℱ* = {*F*_*i*_(*x*)|*i* ∈ 1, …, *N*} and weights *w*, the linear pool is calculated as

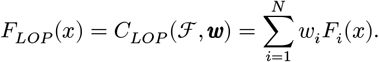

For a set of PMF values, {*f*_*i*_(*x*)|*i* ∈ 1, …, *N*}, the linear pool can be equivalently calculated as 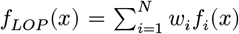. Statistically this amounts to a mixture of the probability distributions, and the resulting probability distribution can be interpreted as one where the constituent probability distributions represent alternative predictions of the future, each of which has a probability *w*_*i*_ of being the true one. For a visual depiction of these equations, see Figure 2 below.

The different averaging methods for probabilistic predictions yield different properties of the resulting ensemble distribution. For example, the variance of the linear pool is 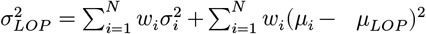, where *μ*_*i*_ is the mean and 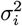 is the variance of individual prediction *i*, and although there is no closed-form variance for the quantile average, the variance of the quantile average will always be less than or equal to that of the linear pool^33^. Both methods generate distributions with the same mean, 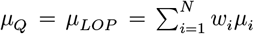, which is the mean of individual model means^33^. The linear pool method preserves variation between individual models, whereas the quantile average cancels away this variation under the assumption it constitutes sampling error^24^.

### 2.3 Applications in public health and infectious disease outbreaks

Multi-model ensembles have become the gold standard for forecasting and prediction efforts that support public health in real time^12,34–36^. One prominent domain is forecasting key characteristics of infectious disease outbreaks, including weekly incidence or healthcare demand over future weeks, disease burden for the entire season, and timing and magnitude of the outbreak peak^14–16,23,37^. Projections of disease outcomes under multiple possible future scenarios have also been used to estimate intervention effectiveness to inform policy^38–40^, and it has been proposed to use short-term forecasts of incidence to inform vaccine efficacy trials^41^. Standard guidelines for reporting of prediction efforts in outbreak and public health settings have also been established^42^.

Across a variety of pathogens and outbreak settings, multi-model ensembles have been shown to produce forecasts that are as good or better than the individual models that compose the ensemble^10,13,14,16–19,23^. Notably, the ensemble does not always outperform the best model, but typically offers improved consistency and robustness over individual models^16,17,20,43^. However, in one instance, a baseline historical average of West Nile Virus cases in the US outperformed most model predictions, including the ensemble^15^.

Examinations of ensemble methodology in infectious disease contexts suggest there is not one method that universally performs best. In short-term forecasting settings, a simple linear pool average of component predictions tends to produce prediction intervals that are too wide (i.e., suggesting outcomes are more uncertain than in reality); beta-transformation^44^ and dynamic weighting^45^ have been suggested to mitigate this problem. A median quantile average has been shown to provide similar performance to a weighted mean in short-term forecasting challenges, while also offering robustness to changes in performance across individuals models^25^, and was thus used as the primary ensemble for short-term forecasts of COVID-19^16,17^. For longer-term predictions of COVID-19, a trimmed LOP ensemble performed best, as models tended to be more overconfident in this setting^18^. The number of models submitting real-time predictions has varied dramatically (from as few as four models for longer-term predictions of COVID-19^18^ to more than 40 for short-term forecasts of COVID-19^16^). Research including applications across influenza and COVID-19 suggests that at least 3 models are needed, with diminishing returns for every model that is added^46^.

The growing body of literature on multi-model ensembles in public health domains emphasizes the utility of these approaches to inform response in real time. Future research on optimizing ensemble performance for different targets and time horizons will further improve utility. Moreover, expanding the use of these methods to other pathogens and countries will enable further methodological development.

## 3 How to implement ensemble calculations

The methods described in Section 2 are implemented via the `hubEnsembles` package in a flexible, easy-to-use framework. Importantly, `hubEnsembles` is situated within the broader hubverse software infrastructure, which provides data standards and conventions for representing and working with model predictions^30^, including for example, collecting and manipulating predictions `(hubUtils)` as well as visualization (`hubVis`). In 2024-2025, the hubverse supported over a dozen collaborative modeling hubs used by public health agencies across the globe. We begin with a short overview of hubverse concepts and conventions that support the process of combining model predictions, supplemented by example predictions provided by the hubverse in `hubExamples`, then explain the implementation of the two primary ensembling functions included in the package, `simple_ensemble()` and `linear_pool()`.

### 3.1 Terminology and data standards in the hubverse

In the hubverse, predictions are always represented in a standardized tabular format called “model output”, codified by the `model_out_tbl` S3 class in `hubUtils` (a package of basic utility functions). Each row represents a single, unique prediction while the columns provide information about what is being predicted, its scope, and value. A single model output object can store and organize many predictions while remaining easy to parse at a glance, which is particularly useful when collecting predictions from multiple models to combine into an ensemble. Any tabular predictions can be transformed into model output using the `as_model_out_tbl` () function from `hubUtils` (see Section 5 for an example).

The `model_out_tbl` class is defined by four standard types of columns: (i) the model ID, which denotes which model has produced the prediction; (ii) the task IDs (also referred to as “task ID variables” or “task ID columns”), which provide details about what is being predicted; (iii) the model output representation, which specify the type of prediction and other identifying information; and (iv) the value of the prediction itself. While most of these columns are always required and have standardized column names, the task ID variables may vary according to the needs of the modeling hub or modeling task^30^.

Table 1 provides an example of model output that stores short-term forecasts of weekly flu hospitalizations for different US states and territories. By reading across the table, we can see that these are quantile predictions (`output_type`) of the quartiles (output type ID: `otid`) from a single model (model_id of “team1-mod”) for four distinct forecast horizons. Here, details about the prediction related to modeling task are represented by the task ID variables loc (location abbreviation), `ref_date` (reference date: the “starting point” of the forecasts), h (horizon: how many weeks into the future, relative to the `ref_date`), and `target` (what is being predicted).

**Table 1:** Example of forecasts for incident influenza hospitalizations, formatted according to hubverse standards. Quantile predictions for the median and 50% prediction intervals from a single model are shown for four distinct horizons. The `output_type_id` column’s name has been shortened to `otid` for brevity. These predictions are a modified subset of the `forecast_outputs` data provided by the `hubExamples` package.

| `model_id` | `loc` | `ref_date` | `h` | `target` | `output_type` | `otid` | `value` |
| --- | --- | --- | --- | --- | --- | --- | --- |
| team1-mod | MA | 2022-12-17 | 0 | wk flu hosp | quantile | 0.25 | 514 |
| team1-mod | MA | 2022-12-17 | 0 | wk flu hosp | quantile | 0.5 | 596 |
| team1-mod | MA | 2022-12-17 | 0 | wk flu hosp | quantile | 0.75 | 713 |
| team1-mod | MA | 2022-12-17 | 1 | wk flu hosp | quantile | 0.25 | 563 |
| team1-mod | MA | 2022-12-17 | 1 | wk flu hosp | quantile | 0.5 | 664 |
| team1-mod | MA | 2022-12-17 | 1 | wk flu hosp | quantile | 0.75 | 803 |
| team1-mod | MA | 2022-12-17 | 2 | wk flu hosp | quantile | 0.25 | 469 |
| team1-mod | MA | 2022-12-17 | 2 | wk flu hosp | quantile | 0.5 | 575 |
| team1-mod | MA | 2022-12-17 | 2 | wk flu hosp | quantile | 0.75 | 705 |
| team1-mod | MA | 2022-12-17 | 3 | wk flu hosp | quantile | 0.25 | 324 |
| team1-mod | MA | 2022-12-17 | 3 | wk flu hosp | quantile | 0.5 | 408 |
| team1-mod | MA | 2022-12-17 | 3 | wk flu hosp | quantile | 0.75 | 512 |

As mentioned previously, task ID variables are not fixed in name, number, or composition to incorporate flexibility in the `model_out_tbl` class. Different modeling efforts may use different sets of task ID columns with different values to define their prediction goals, or may simply choose distinct names to represent the same concept. For example, the date task column was named `ref_date` above but could easily be called `origin_date` or `forecast_date` instead. Some standard examples of task ID variables are available on the hubverse documentation website^30^.

The “model output representation” columns `output_type` and `output_type_id` contain metadata about how the predictions are conveyed. The hubverse data standards require that these columns are included in a `model_out_tbl`. The `output_type` column defines how a prediction is represented and may be `“mean”` or `“median”` for point predictions, or one of `“quantile”`, `“cdf”`, `“pmf”`, or `“sample”` for probabilistic predictions. The `output_type_id` provides additional identifying information for prediction and is specific to the particular `output_type` (see Table 2). The last column, value, always contains the numeric value of the prediction, regardless of output type. Requirements for the values of the `output_type_id` and `value` columns associated with each valid output type are summarized in Table 2.

**Table 2:** A table summarizing how the model output representation columns are used for predictions of different output types; adapted from hubverse documentation^30^. (*Rows of sample predictions from a particular model that share an output type ID value may be assumed to represent a single sample from a joint distribution across multiple levels of the task ID variables.)

| output_type | output_type_id | value |
| --- | --- | --- |
| mean | NA (not used for mean predictions) | Numeric: The mean of the predictive distribution |
| median | NA (not used for median predictions) | Numeric: The median of the predictive distribution |
| quantile | Numeric between 0.0 and 1.0: A quantile level | Numeric: The quantile of the predictive distribution at the quantile level specified by the **output_type_id** |
| cdf | String or numeric naming a possible value of the target variable | Numeric between 0.0 and 1.0: The cumulative probability of the predictive distribution at the value of the outcome variable specified by the **output_type_id** |
| pmf | String naming a possible category of a discrete outcome variable | Numeric between 0.0 and 1.0: The probability mass of the predictive distribution when evaluated at a specified level of a categorical outcome variable |
| sample | Integer or string identifying the sample* | Numeric: A single sample value from the predictive distribution |

All output types can summarize predictions from univariate marginal distributions, e.g. for a single location and time point. The sample output type, which represents randomly drawn values from a probabilistic predictive distribution, is unique in that it can additionally represent predictions from joint predictive distributions. This means that samples may encode dependence across combinations of multiple values for task ID variables, for example across multiple locations and/or time points. In this case, rows of sample predictions with the same index (specified by the `output_type_id`) from a particular model may be assumed to correspond to a single sample from a joint distribution.

For quantile predictions, the `output_type_id` is a numeric value between 0 and 1 specifying the cumulative probability associated with the quantile prediction. In the notation we defined in Section 2, the `output_type_id` corresponds to *θ* and the `value` is the quantile prediction *F* ^−1^(*θ*). For CDF or PMF predictions, the `output_type_id` is the target variable value *x* at which the cumulative distribution function or probability mass function for the predictive distribution should be evaluated, and the value column contains the predicted *F* (*x*) or *f*(*x*), respectively.

The hubverse also provides standards for target data (i.e., observed data corresponding to each prediction target), which can be stored in one of two formats: target time series or oracle output. The two tabular representations differ in terms of columns and purposes. Target time series data is a more traditional representation of the observed “truth” in a time series format with minimal columns; this format usually serves as calibration data for generating forecasts or might be used for visualization of predictions. Oracle output, on the other hand, represents prediction that an “oracle model” would have made had it known the observed values in advance. This format resembles model output data and is suited for evaluating forecasts. Some examples of target data are given in Section 4.

### 3.2 Ensemble functions in hubEnsembles

The `hubEnsembles` package contains two functions that perform ensemble calculations: `simple_ensemble()`, which applies some function to each model prediction, and `linear_pool()`, which computes an ensemble using the linear opinion pool method. In the following sections, we outline the implementation details for each function and how these implementations correspond to the statistical ensembling methods described in Section 2. A short description of the calculation performed by each function is summarized by output type in Table 3.

**Table 3:**
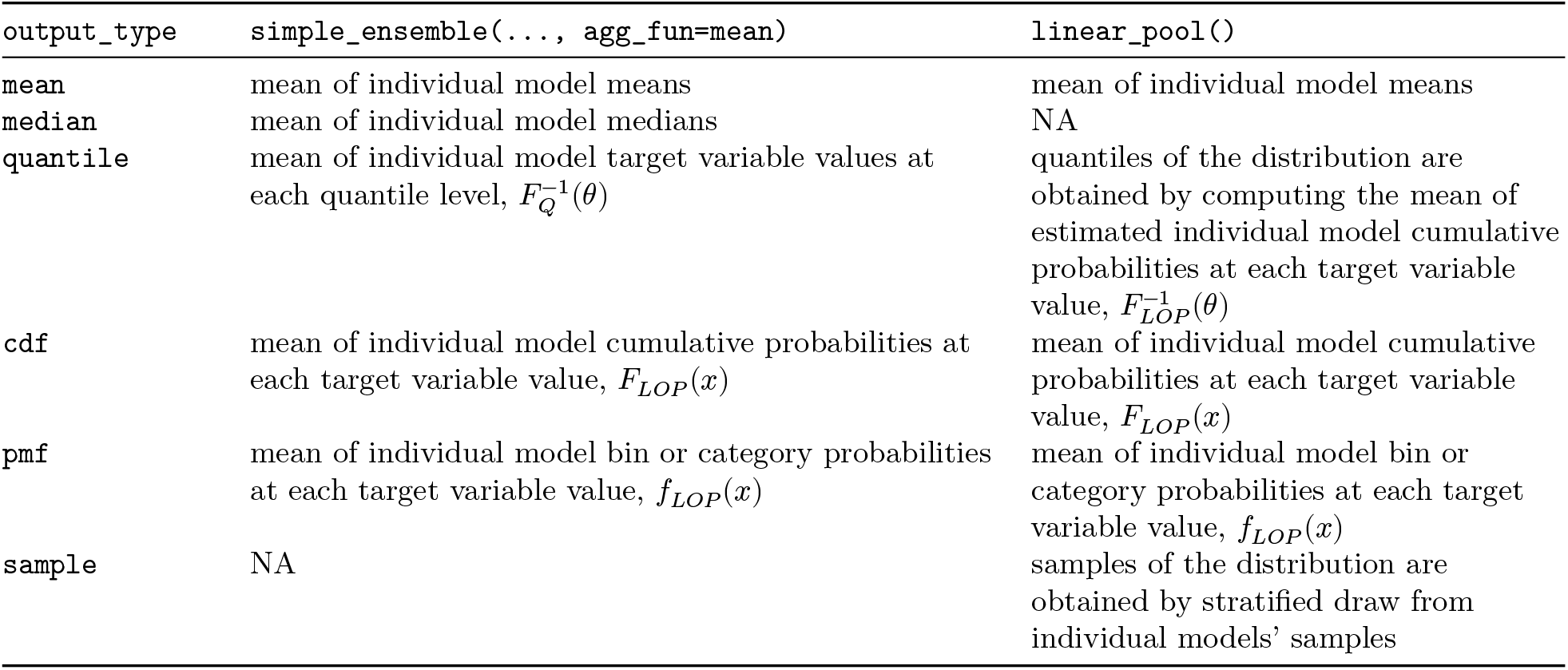
Summary of ensemble function calculations for each output type. The ensemble function determines the operation that is performed, and in the case of probabilistic output types (quantile, CDF, PMF), this also determines what ensemble distribution is generated (quantile average,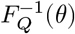, or linear pool, *F*_*LOP*_ (*x*)). The ensembled predictions are returned in the same output type as the inputs. Thus, the output type determines how the resulting ensemble distribution is summarized (as a quantile, *F* ^−1^(*θ*), cumulative probability, *F* (*x*), or probability *f*(*x*)). Estimating individual model cumulative probabilities is required to compute a `linear_pool()` for predictions of `quantile` output type; see Section 3.2.2 on the linear pool operation for details. In the case of `simple_ensemble()`, we show the calculations for the default case where `agg_fun = mean;` however, if another aggregation function is chosen (e.g., `agg_fun = median`), that calculation would be performed instead. For example, `simple_ensemble(…, agg_fun = median)` applied to predictions of mean output type would return the median of individual model means.

#### 3.2.1 Simple ensemble

The `simple_ensemble()` function directly computes an ensemble from component model outputs by combining them via an aggregation function (*C*) within each unique combination of task ID variables, output types, and output type IDs. This function can be used to summarize predictions of output types mean, median, quantile, CDF, and PMF. The mechanics of the ensemble calculations are the same for each of the output types, though the resulting statistical ensembling method differs for different output types (Table 3).

By default, `simple_ensemble()` uses the mean for the aggregation function *C* and equal weights for all models. For point predictions with a mean or median output type, the resulting ensemble prediction is an equally weighted average of the individual models’ predictions. For probabilistic predictions in a quantile format, by default `simple_ensemble()` calculates an equally weighted average of individual model target variable values at each quantile level, which is equivalent to a quantile average. For model outputs in a CDF or PMF format, by default `simple_ensemble()` computes an equally weighted average of individual model (cumulative or bin) probabilities at each target variable value, which is equivalent to the linear pool method.

Any aggregation function *C* may be specified by the user. For example, a median ensemble may also be created by specifying `median` as the aggregation function, or a custom function may be passed to the `agg_fun` argument to create other ensemble types. Similarly, model weights can be specified to create a weighted ensemble.

#### 3.2.2 Linear pool

The `linear_pool()` function implements the linear opinion pool (LOP) method for ensembling predictions. Currently, this function can be used to combine predictions with output types mean, quantile, CDF, PMF, and sample. Unlike `simple_ensemble()`, this function handles its computation differently based on the output type. For the CDF, PMF, and mean output types, the linear pool method is equivalent to calling `simple_ensemble()` with a mean aggregation function (see Table 3), since `simple_ensemble()` produces a linear pool prediction (an average of individual model cumulative or bin probabilities).

For the sample output type, the LOP method collects a stratified draw of the individual models’ predictions and pools them into a single ensemble distribution. By default, all samples are used to create this ensemble. Additionally, only equally-weighted linear pools of samples are supported by the `hubEnsembles` package during this time. Samples may also be converted to another common output type such as quantiles or bin probabilities (as the main scientific interest often concerns a summary of samples), and other ensemble methods may then be utilized for that output type.

For the quantile output type, implementation of LOP is comparatively less straightforward. This is because LOP averages cumulative probabilities at each value of the target variable, but the predictions are given as quantiles (on the scale of the target variable) for fixed quantile levels. The value for these quantile predictions will generally differ between models; hence, we are typically not provided cumulative probabilities at the same values of the target variable for all component predictions. This lack of alignment between cumulative probabilities for the same target variable values impedes computation of LOP from quantile predictions and is illustrated in panel A of Figure 2.

Given that LOP cannot be directly calculated from quantile predictions, we must first obtain an estimate of the CDF for each component distribution from the provided quantiles, combine the CDFs, then calculate the quantiles using the ensemble’s CDF. We perform this calculation in three main steps, assisted by the `distfromq` package^47^ for the first two:

1. Interpolate and extrapolate from the provided quantiles for each component model to obtain an estimate of the CDF of that particular distribution.
2. Draw samples from each component model distribution. To reduce Monte Carlo variability, we use quasi-random samples corresponding to quantiles of the estimated distribution^48^.
3. Pool the samples from all component models and extract the desired quantiles.

For step 1, functionality in the `distfromq` package uses a monotonic cubic spline for interpolation on the interior of the provided quantiles. The user may choose one of several distributions to perform extrapolation of the CDF tails. These include normal, lognormal, and Cauchy distributions, with “normal” set as the default. A location-scale parameterization is used, with separate location and scale parameters chosen in the lower and upper tails so as to match the two most extreme quantiles. The sampling process described in steps 2 and 3 approximates the linear pool calculation described in Section 2.

## 4 A simple demonstration of multi-model ensembles

In this section, we provide a simple example to illustrate how to compute a multi-model ensemble and compare the methods supported by the functions of `hubEnsembles`. In doing so, we use a number of other packages available through the hubverse, including to access example data and to visualize outputs. See the Code Availability Statement for details about implementation and required package versions.

### 4.1 Example data: a forecast hub

The first step in generating a multi-model ensemble is to gather the predictions we wish to combine. In this example, we use some short-term forecasts already formatted as model output data from the `hubExamples` package. These model outputs are from a larger example modeling hub, created using a modified subset of predictions from the FluSight Forecasting challenge (discussed in further detail in Section 5). In addition to toy model output data, the example hub also includes observed data in the form of target time series data and oracle output.

The model output is stored in a data object named `forecast_outputs` and contains mean, median, quantile, and sample forecasts of future incident influenza hospitalizations, as well as CDF and PMF forecasts of hospitalization intensity. Each prediction is described by five task ID variables: the location for which the forecast was made (`location`), the date on which the forecast was made (`reference_date`), the number of steps ahead (`horizon`), the date of the forecast prediction (`target_end_date`, a combination of the date the forecast was made and the forecast horizon), and the forecast target (`target`). We begin by examining the predictions for weekly incident influenza hospitalizations, displaying a subset of each output type in Table 4.

**Table 4:** Example model output for forecasts of weekly incident influenza hospitalizations. A subset of example model output is shown: 1-week ahead forecasts made on 2022-12-17 for Massachusetts from three distinct models; only the mean, median, select samples, and the 5th, 25th, 75th and 95th quantiles are displayed. The `location, reference_date` and `target_end_date` columns have been omitted for brevity. This example data is provided in the `hubExamples` package.

| model_id | target | horizon | output_type | output_type_id | value |
| --- | --- | --- | --- | --- | --- |
| Flusight-baseline | wk inc flu hosp | 1 | mean | NA | 582.07 |
| Flusight-baseline | wk inc flu hosp | 1 | median | NA | 582.00 |
| Flusight-baseline | wk inc flu hosp | 1 | quantile | 0.05 | 496.00 |
| Flusight-baseline | wk inc flu hosp | 1 | quantile | 0.25 | 566.00 |
| Flusight-baseline | wk inc flu hosp | 1 | quantile | 0.75 | 598.00 |
| Flusight-baseline | wk inc flu hosp | 1 | quantile | 0.95 | 668.00 |
| Flusight-baseline | wk inc flu hosp | 1 | sample | 2101 | 606.00 |
| Flusight-baseline | wk inc flu hosp | 1 | sample | 2102 | 576.00 |
| Flusight-baseline | wk inc flu hosp | 1 | sample | 2103 | 578.00 |
| MOBS-GLEAM_FLUH | wk inc flu hosp | 1 | mean | NA | 704.73 |
| MOBS-GLEAM_FLUH | wk inc flu hosp | 1 | median | NA | 664.00 |
| MOBS-GLEAM_FLUH | wk inc flu hosp | 1 | quantile | 0.05 | 446.00 |

**Table 5:** Example target time series data for incident influenza hospitalizations. This table includes target time series data for Massachusetts (FIPS code 25) between 2022-11-01 and 2023-02-01. The target data is provided in the `hubExamples` package.

| date | location | observation |
| --- | --- | --- |
| 2022-11-05 | 25 | 31 |
| 2022-11-12 | 25 | 43 |
| 2022-11-19 | 25 | 79 |
| 2022-11-26 | 25 | 221 |
| 2022-12-03 | 25 | 446 |
| 2022-12-10 | 25 | 578 |
| 2022-12-17 | 25 | 694 |
| 2022-12-24 | 25 | 769 |

| date | location | observation |
| --- | --- | --- |
| 2022-12-31 | 25 | 733 |
| 2023-01-07 | 25 | 466 |
| 2023-01-14 | 25 | 238 |
| 2023-01-21 | 25 | 122 |
| 2023-01-28 | 25 | 71 |

While the `hubExamples` package provides both formats of target data, we focus on the target time series data (Table 5) which is convenient for making forecasts and plotting. The `forecast_target_ts` data object provides observed incident influenza hospitalizations in a given week for given location using columns `observation, date`, and `location`. The forecast-specific task ID variables `reference_date` and `horizon` are not relevant for this time series representation of the target data, and are thus not included as columns.

We can plot the quantile and median forecasts and the target time series data (Figure 3) shown above using the `plot_step_ahead_model_output()` function from `hubVis`, another package in the hubverse suite for visualizing model outputs. We subset the model output data and the target data to the location and time horizons we are interested in.

**Figure 3:**
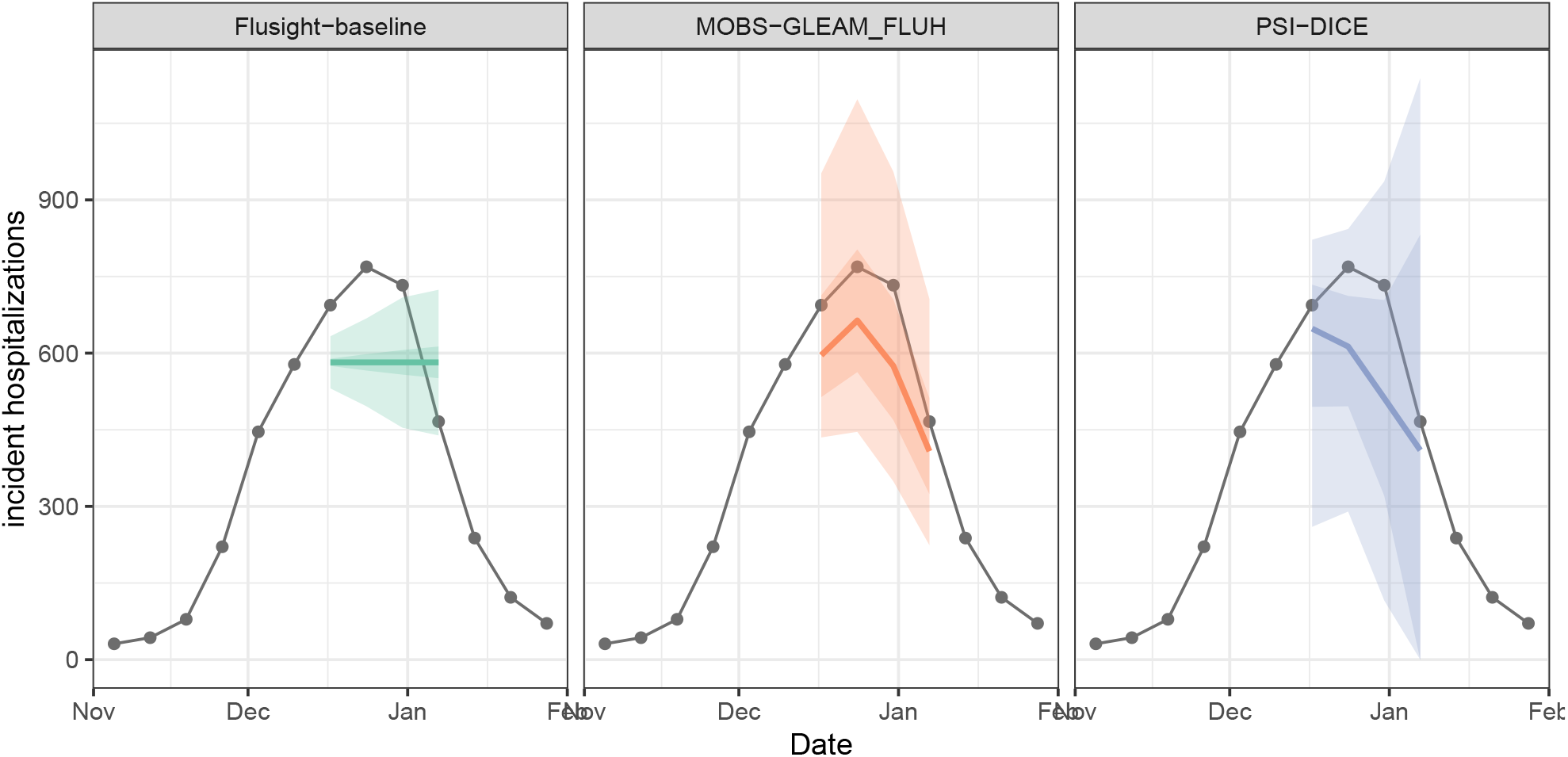
One example set of quantile forecasts for weekly incident influenza hospitalizations in Massachusetts from each of three models (panels). Forecasts are represented by a median (line), 50% and 90% prediction intervals (ribbons). Gray points represent observed incident hospitalizations.

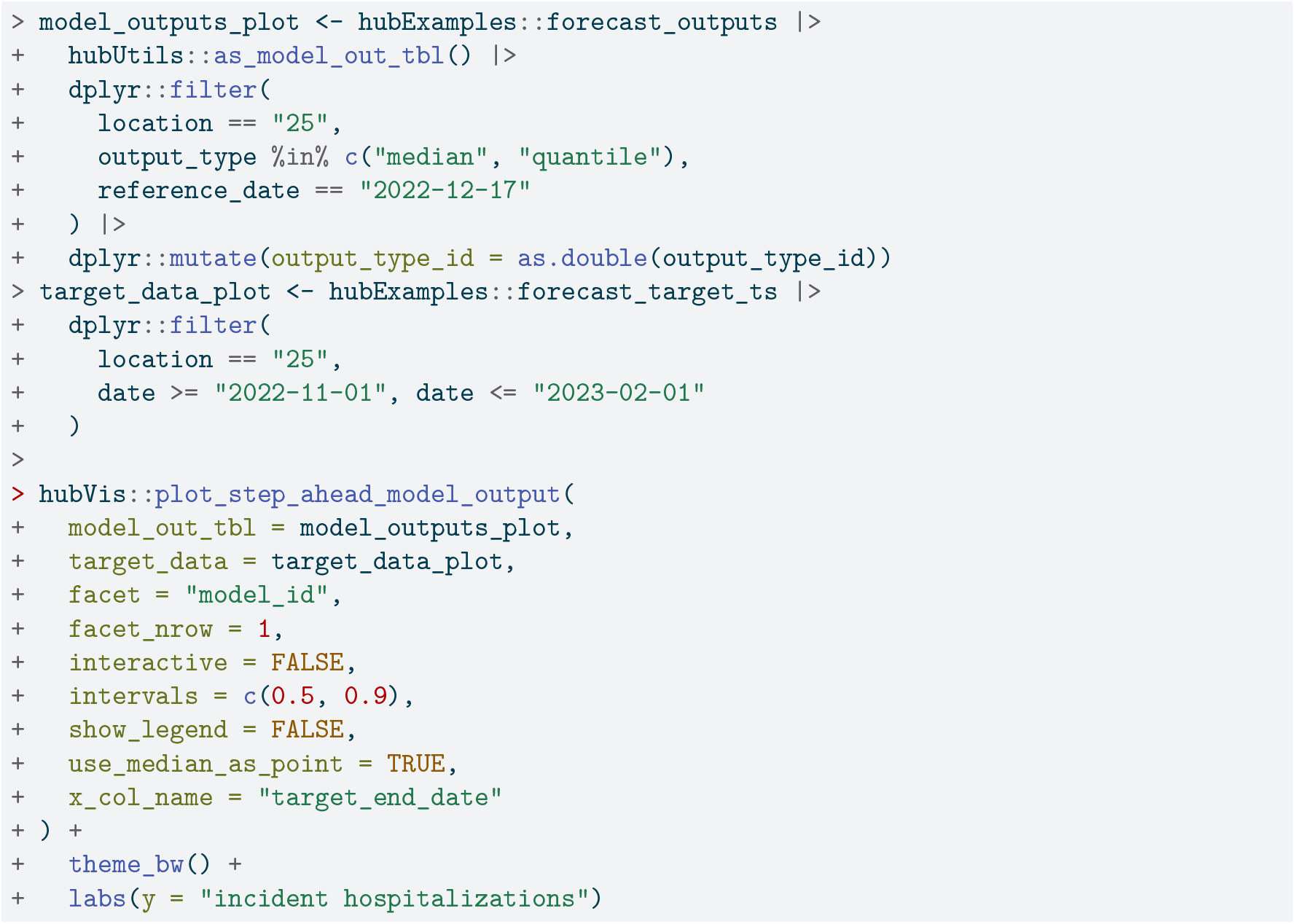

Next, we examine the PMF forecasts for hospitalization intensity in the example model output data. For this target, teams forecasted the probability that hospitalization intensity will be “low”, “moderate”, “high”, or “very high”. These four categories are determined by thresholds for weekly hospital admissions per 100,000 population. In other words, “low” hospitalization intensity in a given week means few incident influenza hospitalizations per 100,000 population are predicted, whereas “very high” hospitalization intensity means many hospitalizations per 100,000 population are predicted. These forecasts are made for the same task ID variables as the `quantile` forecasts of incident hospitalizations except for the target, which is “wk flu hosp rate category” for these categorical predictions.

We show a representative example of the hospitalization intensity category forecasts in Table 6. Because these forecasts are PMF output type, the output_type_id column specifies the bin of hospitalization intensity and the value column provides the forecasted probability of hospitalization incidence being in that category. Values sum to 1 across bins. For the MOBS-GLEAM_FLUH and PSI-DICE models, incidence is forecasted to decrease over the horizon (Figure 3), and correspondingly, there is lower probability of “high” and “very high” hospitalization intensity for later horizons (Figure 4).

**Table 6:** Example PMF model output for forecasts of incident influenza hospitalization intensity. A subset of predictions are shown: 1-week ahead PMF forecasts made on 2022-12-17 for Massachusetts from three distinct models. We round the forecasted probability (in the value column) to two digits. The `location, reference_date` and `target_end_date` columns have been omitted for brevity. This example data is provided in the `hubExamples` package.

| model_id | target | horizon | output_type | output_type_id | value |
| --- | --- | --- | --- | --- | --- |
| Flusight-baseline | wk flu hosp rate category | 1 | pmf | low | 0.000 |
| Flusight-baseline | wk flu hosp rate category | 1 | pmf | moderate | 0.003 |
| Flusight-baseline | wk flu hosp rate category | 1 | pmf | high | 0.073 |
| Flusight-baseline | wk flu hosp rate category | 1 | pmf | very high | 0.924 |
| MOBS-GLEAM_FLUH | wk flu hosp rate category | 1 | pmf | low | 0.000 |
| MOBS-GLEAM_FLUH | wk flu hosp rate category | 1 | pmf | moderate | 0.002 |
| MOBS-GLEAM_FLUH | wk flu hosp rate category | 1 | pmf | high | 0.163 |
| MOBS-GLEAM_FLUH | wk flu hosp rate category | 1 | pmf | very high | 0.835 |
| PSI-DICE | wk flu hosp rate category | 1 | pmf | low | 0.013 |

Table 6: Example PMF model output for forecasts of incident influenza hospitalization intensity.
| model_id | target | horizon | output_type | output_type_id | value |
| --- | --- | --- | --- | --- | --- |
| PSI-DICE | wk flu hosp rate category | 1 | pmf | moderate | 0.065 |
| PSI-DICE | wk flu hosp rate category | 1 | pmf | high | 0.218 |
| PSI-DICE | wk flu hosp rate category | 1 | pmf | very high | 0.704 |

**Figure 4:**
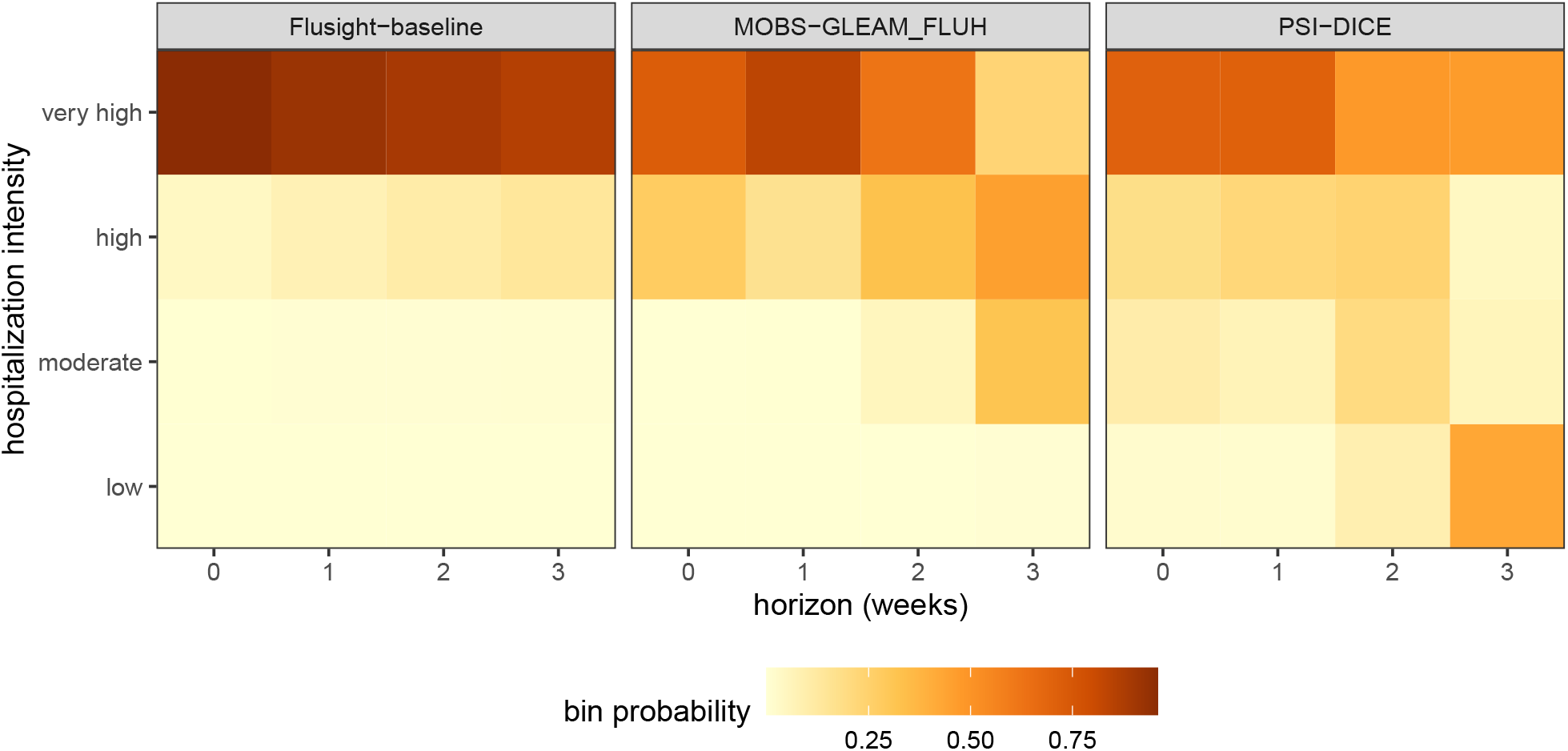
One example PMF forecast of incident influenza hospitalization intensity is shown for each of three models (panels). Each cell shows the forecasted probability of a given hospitalization intensity bin (low, moderate, high, and very high) for each forecast horizon (0-3 weeks ahead). Darker colors indicate higher forecasted probability.

### 4.2 Creating ensembles with simple_ensemble

Using the default options for `simple_ensemble()`, we can generate an equally weighted mean ensemble for each unique combination of values for the task ID variables, the `output_type` and the `output_type_id`. Recall that this function corresponds to different statistical ensemble methods for different output types: for the quantile output type in our example data, the resulting ensemble is a quantile average, while for the CDF and PMF output types, the ensemble is a linear pool (Table 3).

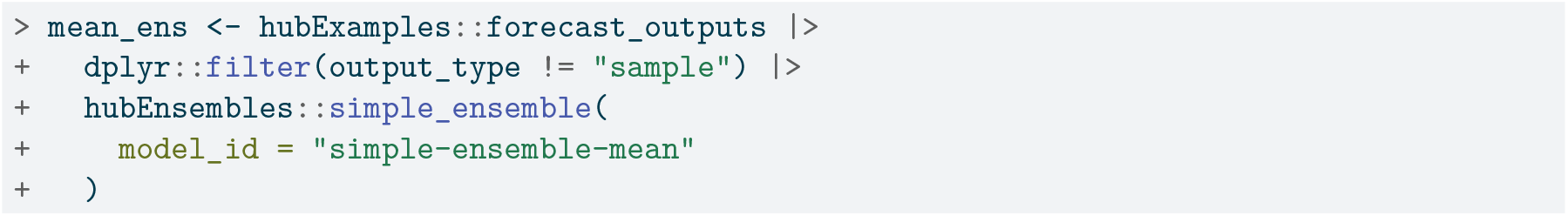

The resulting model output has the same structure as the original model output data (Table 7), with columns for model ID, task ID variables, output type, output type ID, and value. We also use model_id = `“simple-ensemble-mean”` to change the name of this ensemble in the resulting model output; if not specified, the default is “hub-ensemble”.

**Table 7:**
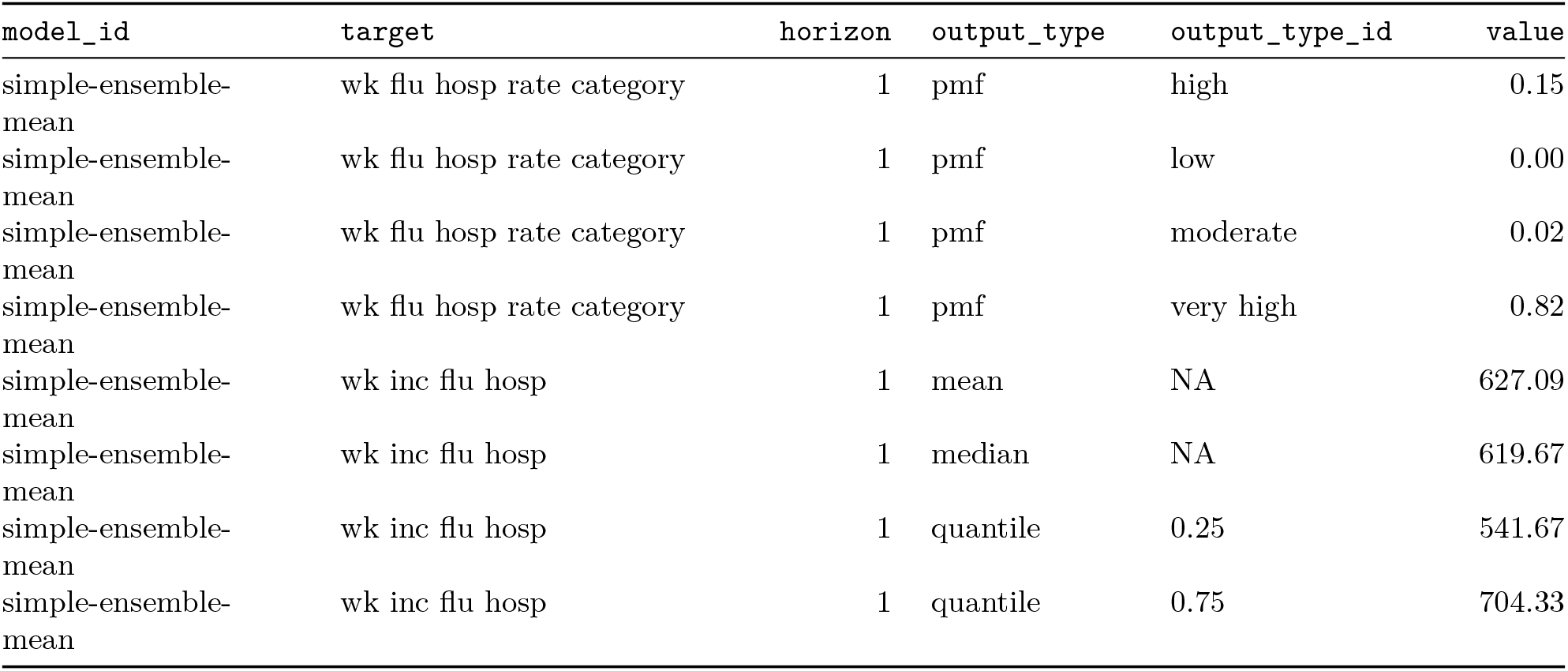
Mean ensemble model output. The values in the model_id column are set by the argument `simple_ensemble(…, model_id)`. Results are generated for all output types, but only a subset are shown: 1-week ahead forecasts made on 2022-12-17 for Massachusetts, with only the mean, median, 25th and 75th quantilesfor the quantile output type and all bins for the PMF output type. The `location, reference_date` and `target_end_date` columns have been omitted for brevity, and the value column is rounded to two digits.

#### 4.2.1 Changing the aggregation function

We can change the function that is used to aggregate model outputs. For example, we may want to calculate a median of the component models’ submitted values for each quantile. We do so by specifying `agg_fun = median`.

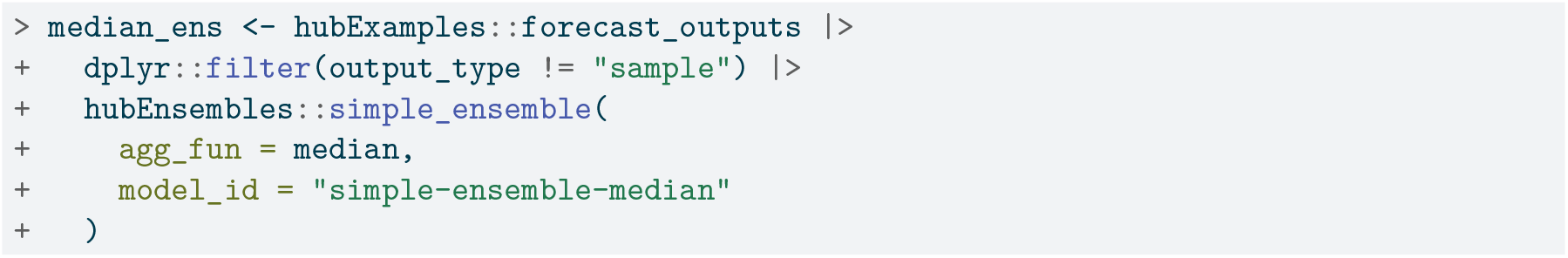

Custom functions can also be passed into the `agg_fun` argument. We illustrate this by defining a custom function to compute the ensemble prediction as a geometric mean of the component model predictions. Any custom function to be used must have an argument x for the vector of numeric values to summarize, and if relevant, an argument w of numeric weights.

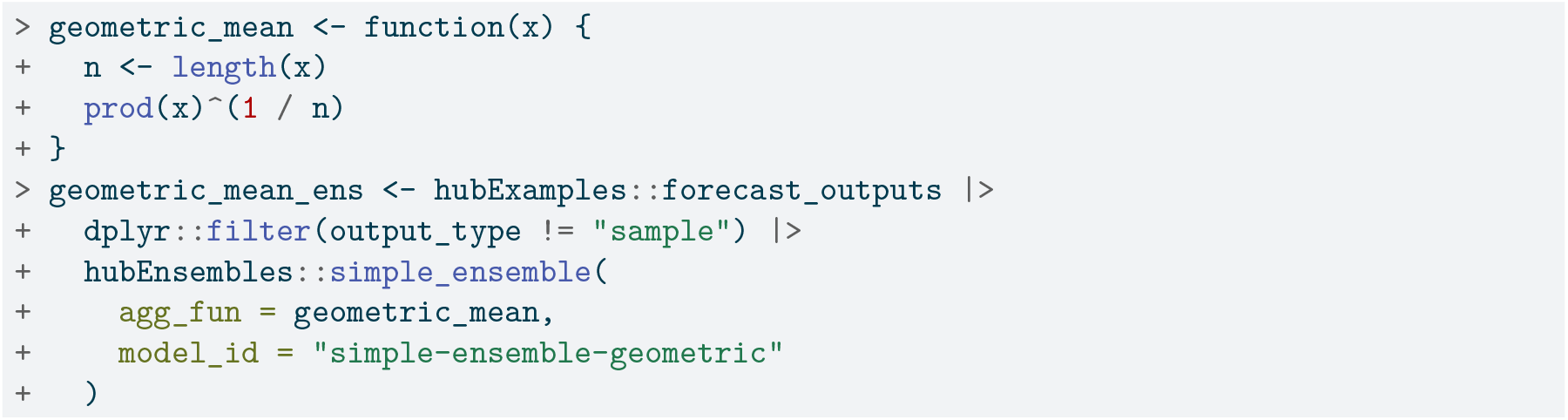

As expected, the mean, median, and geometric mean each give us slightly different resulting ensembles. The median point estimates, 50% prediction intervals, and 90% prediction intervals in Figure 5 demonstrate this.

**Figure 5:**
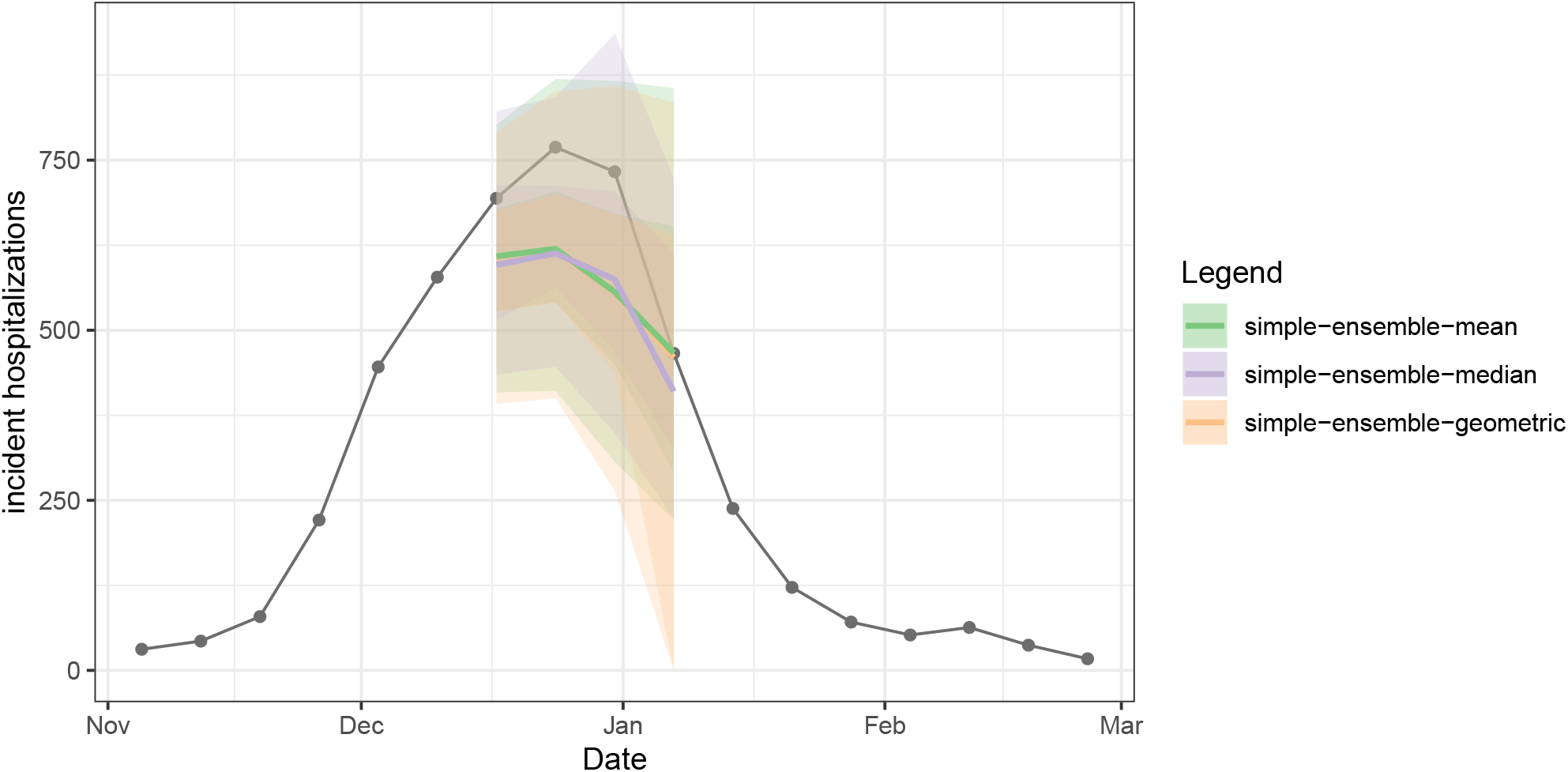
Three different ensembles for weekly incident influenza hospitalizations in Massachusetts. Each ensemble combines individual predictions from the example hub (Figure 3) using a different method: arithmetic mean, geometric mean, or median. All methods correspond to variations of the quantile average approach. Ensembles are represented by a median (line), 50% and 90% prediction intervals (ribbons). Geometric mean ensemble and simple mean ensemble generate similar estimates in this case.

#### 4.2.2 Weighting model contributions

We can weight the contributions of each model in the ensemble using the `weights` argument of `simple_ensemble()`. This argument takes a `data.frame` that should include a `model_id` column containing each unique model ID and a `weight` column. In the following example, we include the baseline model in the ensemble, but give it less weight than the other forecasts.

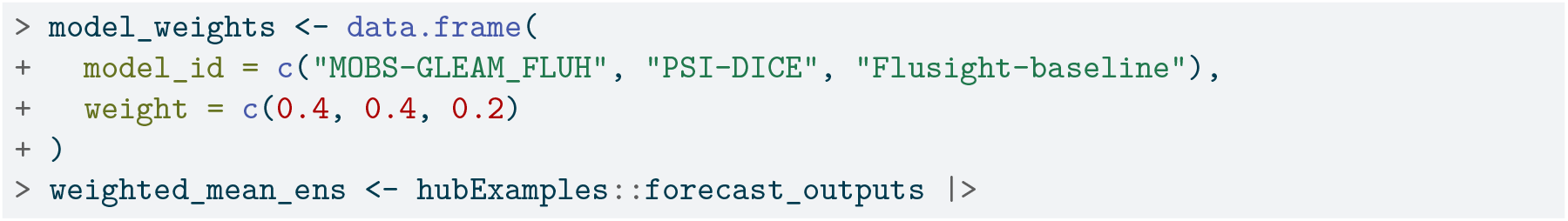
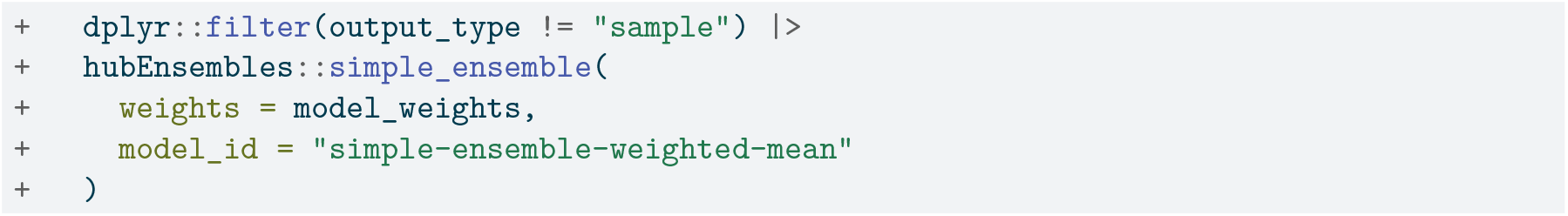

### 4.3 Creating ensembles with linear_pool

We can also generate a linear pool ensemble, or distributional mixture, using the `linear_pool()` function; this function can be applied to predictions with an `output_type` of mean, quantile, sample, CDF, or PMF. Our example hub includes the median output type, so we exclude it from the calculation.

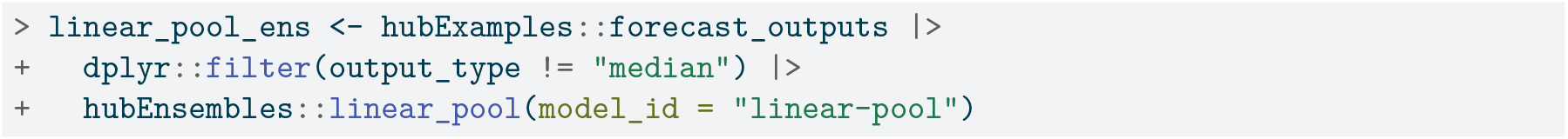

As described above, for `quantile` model outputs, the `linear_pool` function approximates the full probability distribution for each component prediction using the value-quantile pairs provided by that model, and then obtains quasi-random samples from that distributional estimate. The number of samples drawn from the distribution of each component model defaults to `1e4`, but this can be changed using the `n_samples` argument.

In Figure 6, we compare ensemble results generated by `simple_ensemble()` and `linear_pool()` for model outputs of output types PMF and quantile. As expected, the results from the two functions are equivalent for the PMF output type: for this output type, the `simple_ensemble()` method averages the predicted probability of each category across the component models, which is the definition of the linear pool ensemble method. This is not the case for the quantile output type, because the `simple_ensemble()` is computing a quantile average.

**Figure 6:**
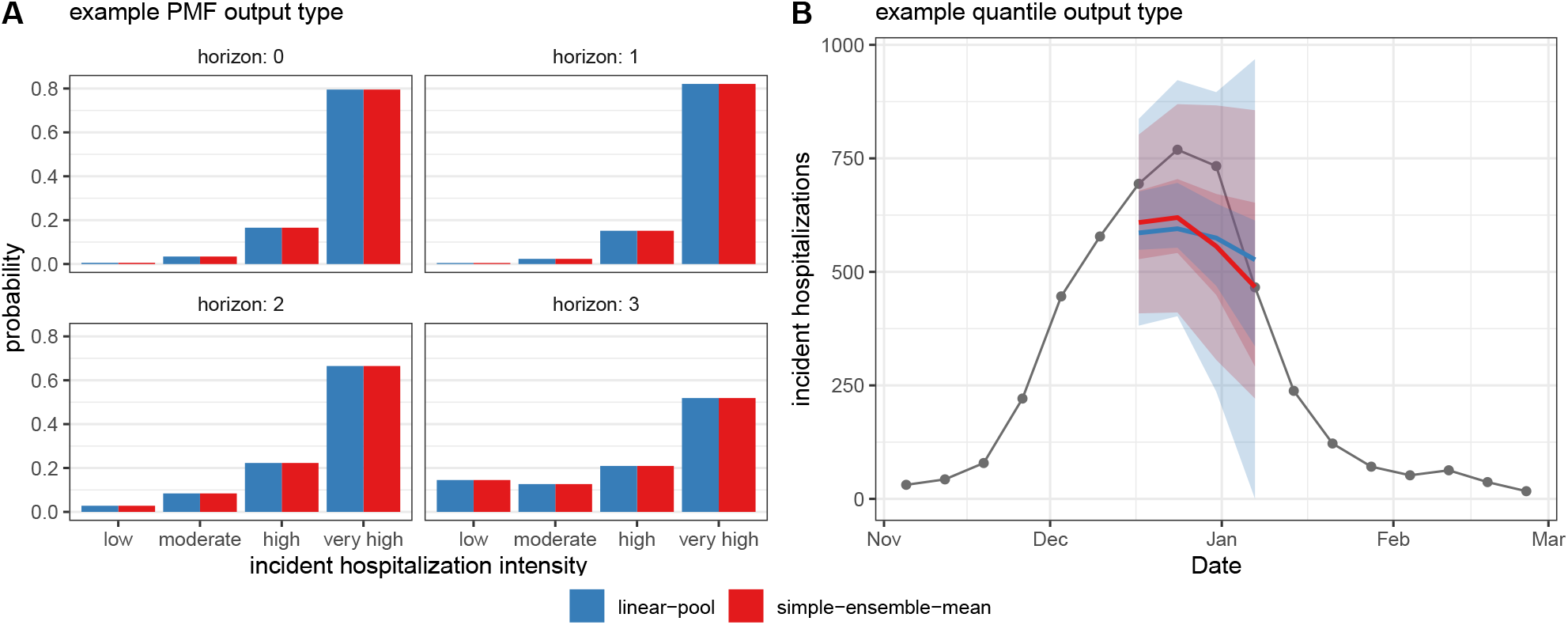
Comparison of results from `linear_pool()` (blue) and `simple_ensemble()` (red). (Panel A) Ensemble predictions of Massachusetts incident influenza hospitalization intensity (classified as low, moderate, high, or very high), which provide an example of PMF output type. (Panel B) Ensemble predictions of weekly incident influenza hospitalizations in Massachusetts, which provide an example of quantile output type. Note, for quantile output type, `simple_ensemble()` corresponds to a quantile average. Ensembles combine individual models from the example hub, and are represented by a median (line), 50% and 90% prediction intervals (ribbons) (Figure 3).

#### 4.3.1 Weighting model contributions

Like with `simple_ensemble()`, we can change the default function settings. For example, weights that determine a model’s contribution to the resulting ensemble may be provided. (Note that we must exclude the sample output type here because it cannot yet be combined into weighted ensembles.)

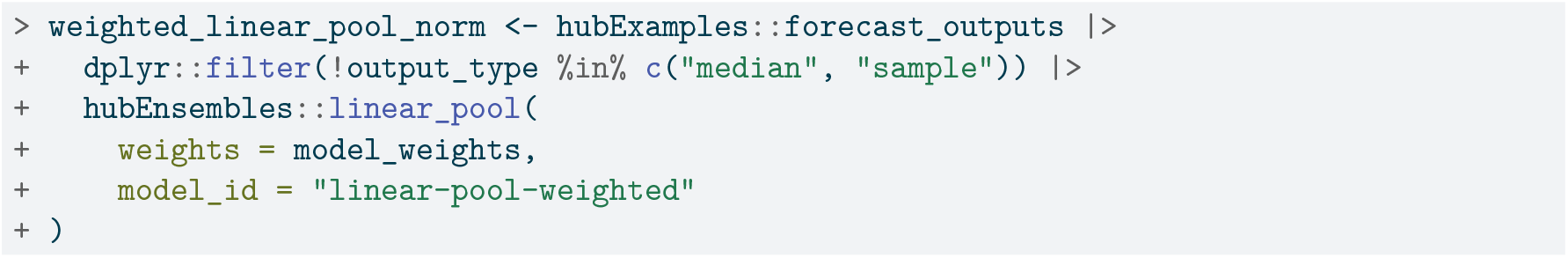

#### 4.3.2 Changing the parametric family used for extrapolation into distribution tails

When requesting a linear pool of quantiles, we may also change the distribution that `distfromq` uses to approximate the tails of component models’ predictive distributions to either log normal or Cauchy using the `tail_dist` argument (the default is normal)^47^. This choice usually does not have a large impact on the resulting ensemble distribution, though, and can only be seen in its outer edges. (For more details and other function options, see the documentation in the `distfromq` package at https://reichlab.io/distfromq/.)

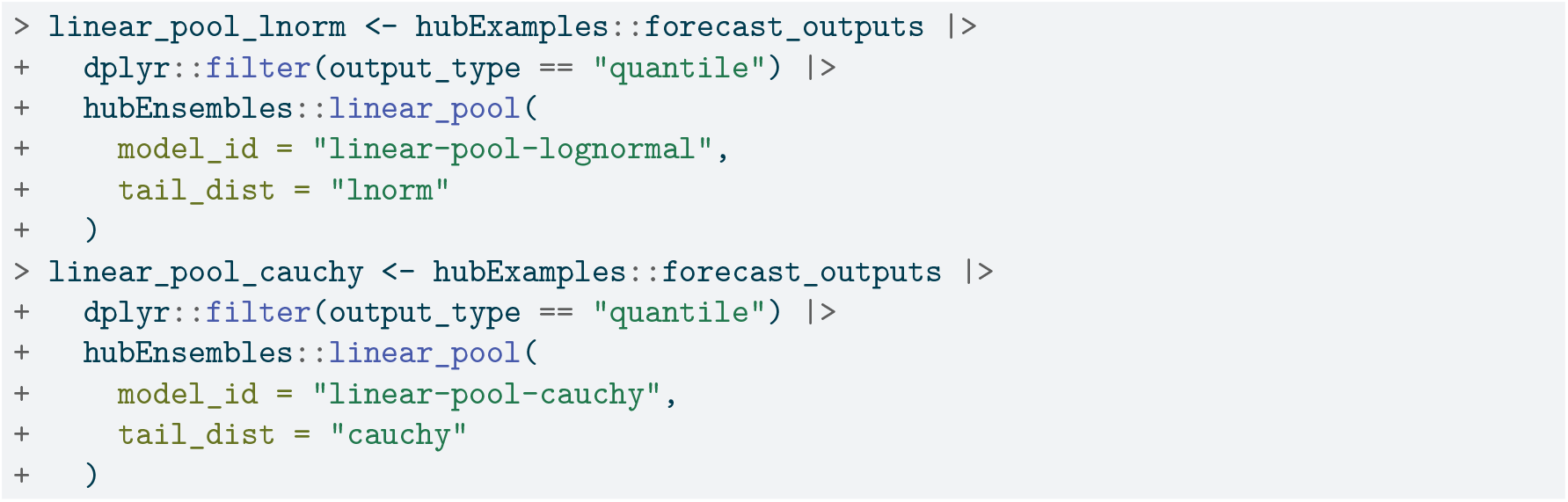

#### 4.3.3 Requesting a subset of input sample predictions to be ensembled

Recall that for the sample output type, `linear_pool()` defaults to creating an equally-weighted ensemble by collecting and returning all provided sample predictions, so that the total number of samples for the ensemble is equal to the sum of the number of samples from all individual models. To change this behavior, the user may instead specify a number of sample predictions for the ensemble to return using the `n_output_samples` argument. Then, a random subset of predictions from individual models will be selected to construct a LOP of samples so that all component models are represented equally. This random selection of samples is stratified by model to ensure approximately the same number of samples from each individual model is included in the ensemble.

When requesting a linear pool composed of a subset of the input sample predictions, the user must identify the task ID variables which together identify a single modeled unit. This group of independent task ID variables is called the compound task ID set and is specified using the `compound_taskid_set` parameter to ensure the subsetting of sample predictions is performed correctly. Samples summarizing a marginal distribution will have a compound task ID set made up of all the task ID variables. On the other hand, samples summarizing a joint distribution will have a compound task ID set that contains only task ID variables for which the joint distribution does not capture dependence. For example, if a joint distribution is estimated across multiple forecast horizons separately for each location, location would be included in the compound task ID set but horizon would not.

Derived task IDs are another subgroup of task ID variables that must be specified in a call to `linear_pool()` for a subsetted sample ensemble; their values are derived from a combination of the values from other task ID variables (which may or may not be part of the compound task ID set). A common example of a derived task ID variable is the target date for a prediction, which is a deterministic function of the reference date of the prediction and the prediction horizon. Generally, the derived task IDs won’t be included in the compound task ID set because they are not needed to identify a single modeled unit for an outcome of interest, *unless* all of the task ID variables their values depend on are already a part of the compound task ID set.

Not all model outputs will contain derived task IDs, in which case the argument may be set to NULL (the default value). However, it is important to provide the `linear_pool()` function with any derived task IDs when calculating an ensemble of (subsetted) samples, as they are used to check that the provided compound task ID set is compatible with the input predictions and the resulting LOP is valid.

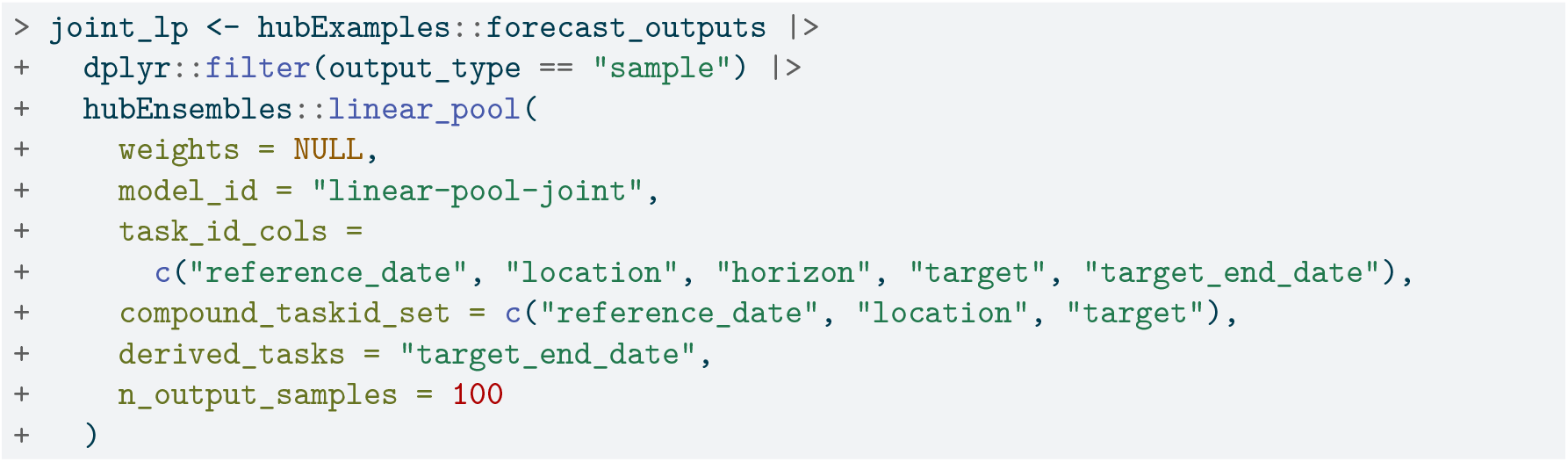

## 5 Example: in-depth analysis of forecast data

To further demonstrate the differences between the two ensemble functions and the utility of the `hubEnsembles` package, we provide a more complex example that walks through the full process of generating multi-model ensembles. This case study gathers real forecasts collected by a modeling hub to create four equally-weighted ensembles, then evaluates their performance to determine the best approach for the application.

The predictions we use to create the ensemble models are sourced from two seasons of the FluSight forecasting challenge. Since 2013, the US Centers for Disease Control and Prevention (CDC) has been soliciting short-term forecasts of seasonal influenza from modeling teams through this collaborative challenge^49^. Using `simple_ensemble()` and `linear_pool()`, we build four equally-weighted, multi-model ensembles to predict weekly influenza hospitalizations: a quantile (arithmetic) mean, a quantile median, a linear pool with normal tails, and a linear pool with lognormal tails. Then, we compare the resulting ensembles’ performance through plotting and scoring their forecasts.

Only a select portion of the code used in this analysis is shown for brevity, but all the functions and scripts used to generate the case study results can be found in the associated GitHub repository (https://github.com/hubverse-org/hubEnsemblesManuscript). In particular, the figures and tables supporting this analysis are generated reproducibly using data from `.rds` files stored in the analysis/data/raw-data directory and scripts in the `inst` directory of the repository.

### 5.1 Data and Methods

We collect the predictions used to generate the four ensembles by querying them from Zoltar^50^, a repository designed to archive forecasts created by the Reich Lab at UMass Amherst. For this analysis we only consider FluSight forecasts in a quantile format from the 2021-2022 and 2022-2023 influenza seasons. These quantile forecasts are stored in two data objects, split by season, called `flu_forecasts-zoltar_21-22.rds` and `flu_forecasts-zoltar_22-23.rds`, which are then joined together into a single data frame. A subset is shown below in Table 8.

**Table 8:** An example prediction of weekly incident influenza hospitalizations pulled directly from Zoltar. The example forecasts were made on May 15, 2023 for California at the 1 week ahead horizon. The forecasts were generated during the FluSight forecasting challenge, then formatted according to Zoltar standards for storage. The `timezero, season, unit, param1, param2`, and `param3` columns have been omitted for brevity. (The `season` column has a value of ‘2021-2022’ or ‘2022-2023’ while the last three ‘param’ columns always have a value of NA.)

| model | target | class | value | cat | prob | sample | quantile | family |
| --- | --- | --- | --- | --- | --- | --- | --- | --- |
| UMass-trends_ensemble | 1 wk ahead inc flu hosp | quantile | 12 | NA | NA | NA | 0.025 | NA |
| UMass-trends_ensemble | 1 wk ahead inc flu hosp | quantile | 17 | NA | NA | NA | 0.100 | NA |
| UMass-trends_ensemble | 1 wk ahead inc flu hosp | quantile | 25 | NA | NA | NA | 0.250 | NA |
| UMass-trends_ensemble | 1 wk ahead inc flu hosp | quantile | 46 | NA | NA | NA | 0.750 | NA |
| UMass-trends_ensemble | 1 wk ahead inc flu hosp | quantile | 56 | NA | NA | NA | 0.900 | NA |
| UMass-trends_ensemble | 1 wk ahead inc flu hosp | quantile | 68 | NA | NA | NA | 0.975 | NA |

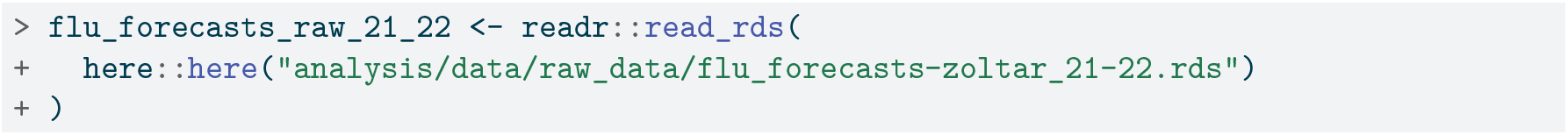
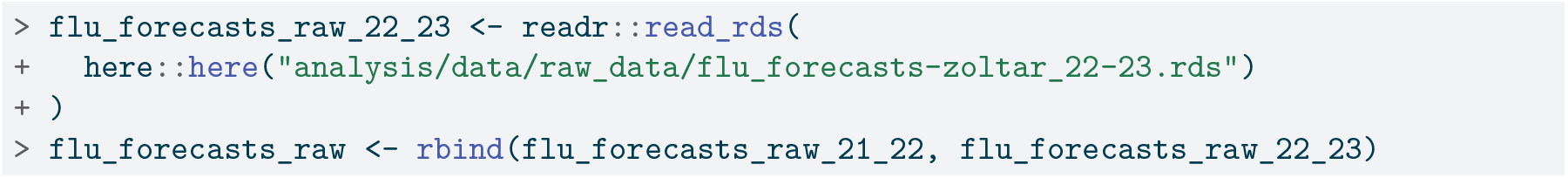

Although these forecasts are in a tabular format, they are not model_out_tbl objects and thus cannot yet be fed into either of the `hubEnsembles` functions. Thus, we must use the `as_model_out_tbl()`^1^ function from `hubUtils` to transform the raw forecasts so that they conform to hubverse standards. Below, we specify the appropriate column mappings in the call with task ID variables of `forecast_date` (when the forecast was made), location, horizon, and `target`.

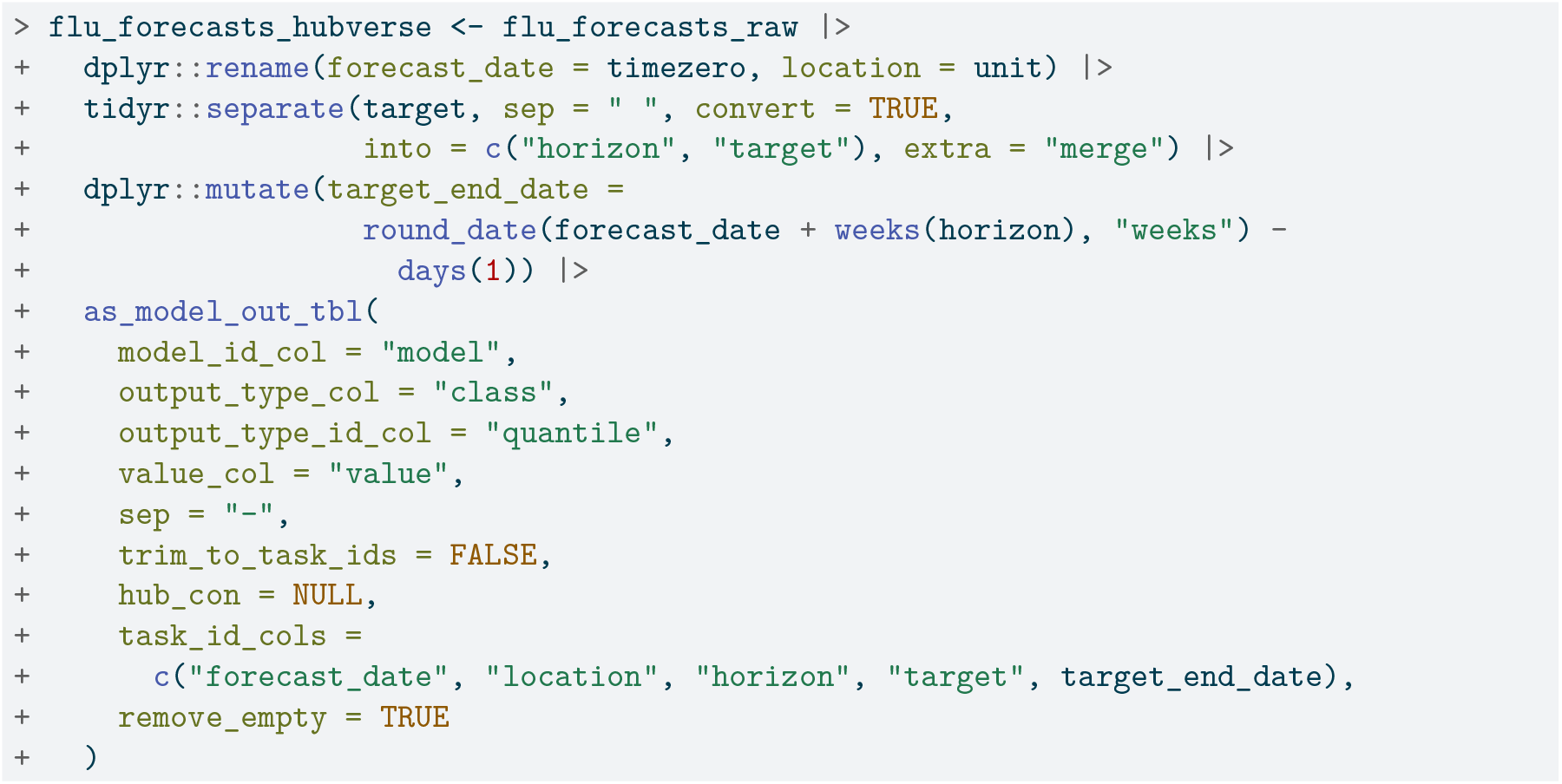

To ensure the quantile mean and median ensemble had consistent component forecast make-up at every quantile level, we only included predictions (defined by a unique combination of task ID variables) that contained all 23 quantiles specified by FluSight (*θ* ∈ {.010, 0.025, .050, .100, …, .900, .950, .990}). This requirement required no further action on our part, since it was consistent with FluSight submission guidelines. However, we did remove the baseline and median ensemble models generated by the FluSight hub from the component forecasts, a choice motivated by the desire to match the composition of models in the official FluSight ensemble.

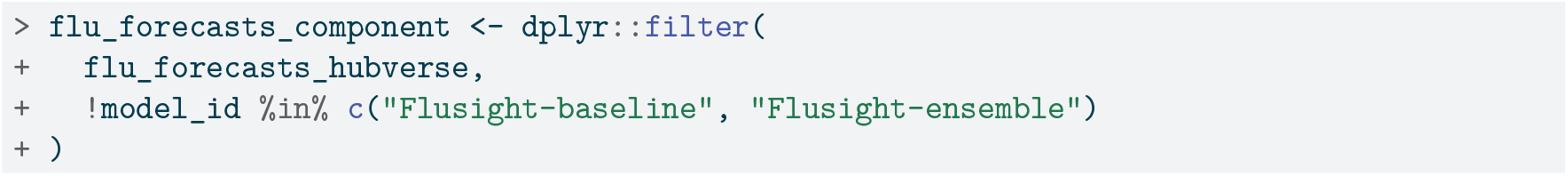

With these inclusion criteria, the final data set of component forecasts consists of predictions from 25 modeling teams and 42 distinct models, 53 forecast dates (one per week), 54 US locations, 4 horizons, 1 target, and 23 quantiles. In the 2021-2022 season, 25 models made predictions for 22 weeks spanning from late January 2022 to late June 2022, and in the 2022-2023 season, there were 31 models making predictions for 31 weeks spanning mid-October 2022 to mid-May 2023. Fourteen of the 42 total models made forecasts for both seasons. Locations consist of the 50 US states, Washington DC, Puerto Rico, the Virgin Islands, and the entire US; horizons 1 to 4 weeks ahead, quantiles the 23 described above, and target week ahead incident flu hospitalization. The values for the forecasts are always non-negative. In Table 9, we provide an example of these predictions, showing select quantiles from a single model, forecast date, horizon, and location.

**Table 9:** An example prediction of weekly incident influenza hospitalizations. The example model output was made on May 15, 2023 for California at the 1 week ahead horizon. The forecast was generated during the FluSight forecasting challenge, then formatted according to hubverse standards post hoc. The `location, forecast_date,` and `season` columns have been omitted for brevity; quantiles representing the endpoints of the central 50%, 80% and 95% prediction intervals are shown.

| model_id | target | horizon | output_type | output_type_id | value |
| --- | --- | --- | --- | --- | --- |
| UMass-trends_ensemble | wk ahead inc flu hosp | 1 | quantile | 0.025 | 12 |
| UMass-trends_ensemble | wk ahead inc flu hosp | 1 | quantile | 0.100 | 17 |
| UMass-trends_ensemble | wk ahead inc flu hosp | 1 | quantile | 0.250 | 25 |
| UMass-trends_ensemble | wk ahead inc flu hosp | 1 | quantile | 0.750 | 46 |
| UMass-trends_ensemble | wk ahead inc flu hosp | 1 | quantile | 0.900 | 56 |
| UMass-trends_ensemble | wk ahead inc flu hosp | 1 | quantile | 0.975 | 68 |

Next, we can combine the predictions into a single `model_out_tbl` object used to generate forecasts for each ensemble method. Then, we call the appropriate function `hubEnsembles` to generate predictions for each equally-weighted ensemble, storing the results in four separate objects of model output data.

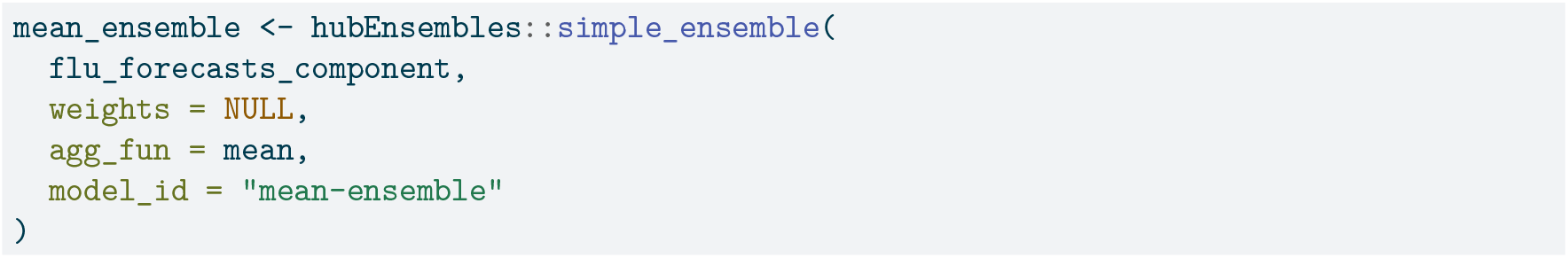
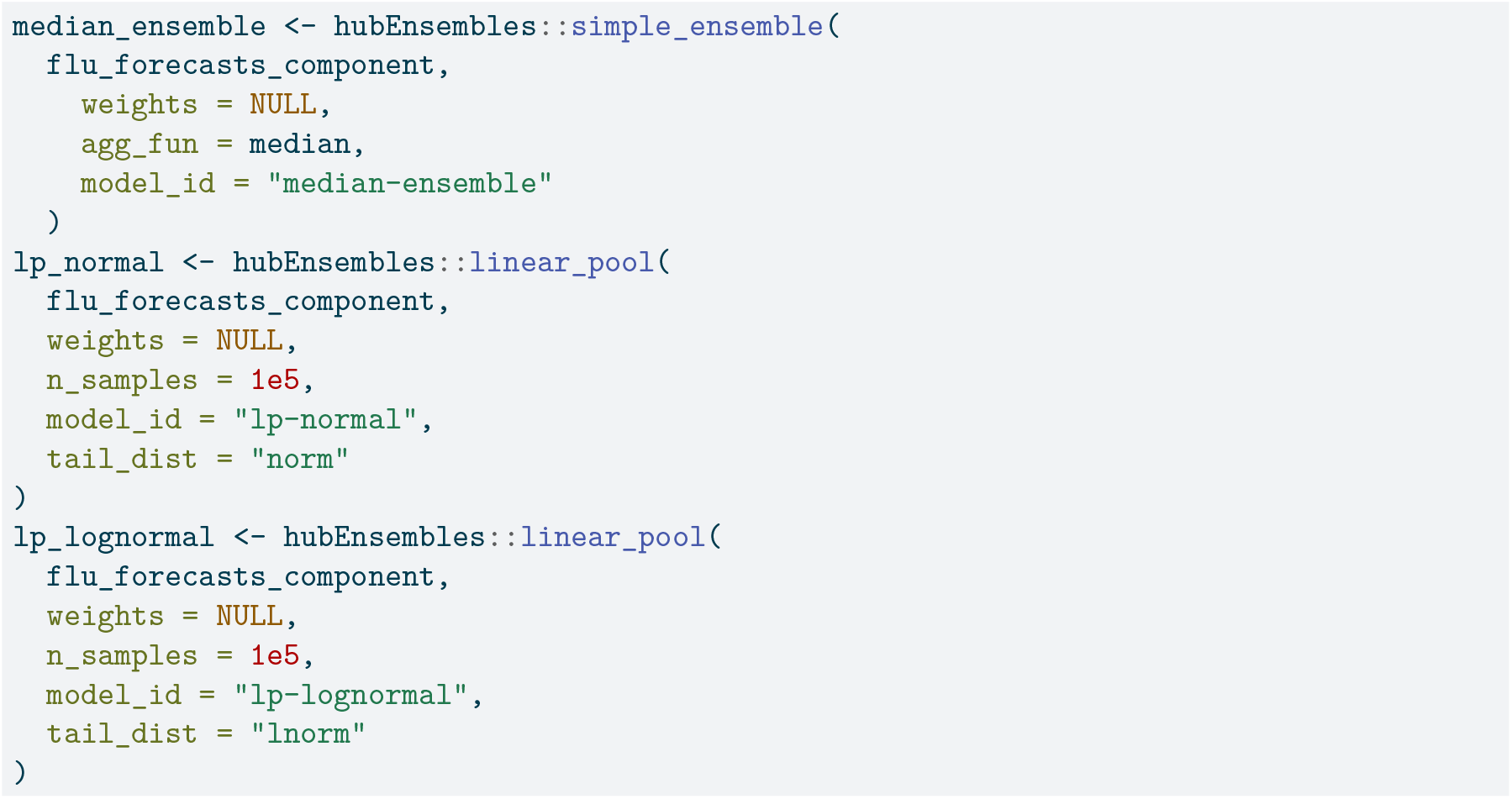

We then evaluate the performance of the ensembles using scoring metrics that measure the accuracy and calibration of their forecasts. We chose several common metrics in forecast evaluation, including mean absolute error (MAE), weighted interval score (WIS)^51^, 50% prediction interval (PI) coverage, and 95% PI coverage. MAE measures the average absolute error of a set of point forecasts; smaller values of MAE indicate better forecast accuracy. WIS is a generalization of MAE for probabilistic forecasts and is an alternative to other common proper scoring rules which cannot be evaluated directly for quantile forecasts^51^. WIS is made up of three component penalties: (1) for over-prediction, (2) for under-prediction, and (3) for the spread of each interval (where an interval is defined by a symmetric set of two quantiles). This metric is a weighted sum of these penalties across all prediction intervals provided. A lower WIS value indicates a more accurate forecast^51^. PI coverage provides information about whether a forecast has accurately characterized its uncertainty about future observations. The 50% PI coverage rate measures the proportion of the time that 50% prediction intervals at that nominal level included the observed value; the 95% PI coverage rate is defined similarly. Achieving approximately nominal (50% or 95%) coverage indicates a well-calibrated forecast.

We also use relative versions of WIS and MAE (rWIS and rMAE, respectively) to understand how the ensemble performance compares to that of the FluSight baseline model. These metrics are calculated as

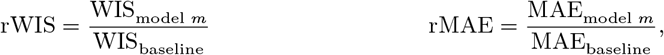

where model *m* is any given model being compared against the baseline. For both of these metrics, a value less than one indicates better performance compared to the baseline while a value greater than one indicates worse performance. By definition, the FluSight baseline itself will always have a value of one for both of these metrics.

Each unique prediction from an ensemble model is scored against target data using the `score_forecasts()`^2^ function from the `covidHubUtils` package, made for scoring tabular infectious disease data with commonly used evaluation metrics including those mentioned above. We use median forecasts taken from the 0.5 quantile for the MAE evaluation.

### 5.2 Performance results across ensembles

The quantile median ensemble has the best overall performance in terms of WIS and MAE (and the relative versions of these metrics), and has coverage rates that were close to the nominal levels (Table 10). The two linear opinion pools have very similar performance to each other. These methods have the second-best performance as measured by WIS and MAE, but they have the highest 50% and 95% coverage rates, with empirical coverage that was well above the nominal coverage rate. The quantile mean performs the worst of the ensembles with the highest MAE, which is substantially different from that of the other ensembles.

**Table 10:** Summary of overall model performance across both seasons, averaged over all locations except the US national location and sorted by ascending WIS. The quantile median ensemble has the best value for every metric except 50% coverage rate, though metric values are often quite similar among the models.

| model | wis | rwis | mae | rmae | cov50 | cov95 |
| --- | --- | --- | --- | --- | --- | --- |
| median-ensemble | 18.158 | 0.794 | 27.360 | 0.933 | 0.597 | 0.922 |
| lp-normal | 19.745 | 0.863 | 27.932 | 0.953 | 0.709 | 0.990 |
| lp-lognormal | 19.747 | 0.863 | 27.933 | 0.953 | 0.708 | 0.990 |
| mean-ensemble | 20.180 | 0.882 | 29.582 | 1.009 | 0.595 | 0.889 |
| Flusight-baseline | 22.876 | 1.000 | 29.315 | 1.000 | 0.604 | 0.881 |

Plots of the models’ forecasts can aid our understanding about the origin of these accuracy differences. For example, the linear opinion pools consistently have some of the widest prediction intervals, and consequently the highest coverage rates. The median ensemble, which has the best WIS, balanced interval width with calibration best overall, with narrower intervals than the linear pools that still achieved near-nominal coverage on average across all time points. The quantile mean’s interval widths vary, though it usually has narrower intervals than the linear pools. However, this model’s point forecasts have a larger error margin compared to the other ensembles, especially at longer horizons. This pattern is demonstrated in Figure 7 for the 4-week ahead forecast in California following the 2022-23 season peak on December 5, 2022. Here, the quantile mean predicted a continued increase in hospitalizations, at a steeper slope than the other ensemble methods.

**Figure 7:**
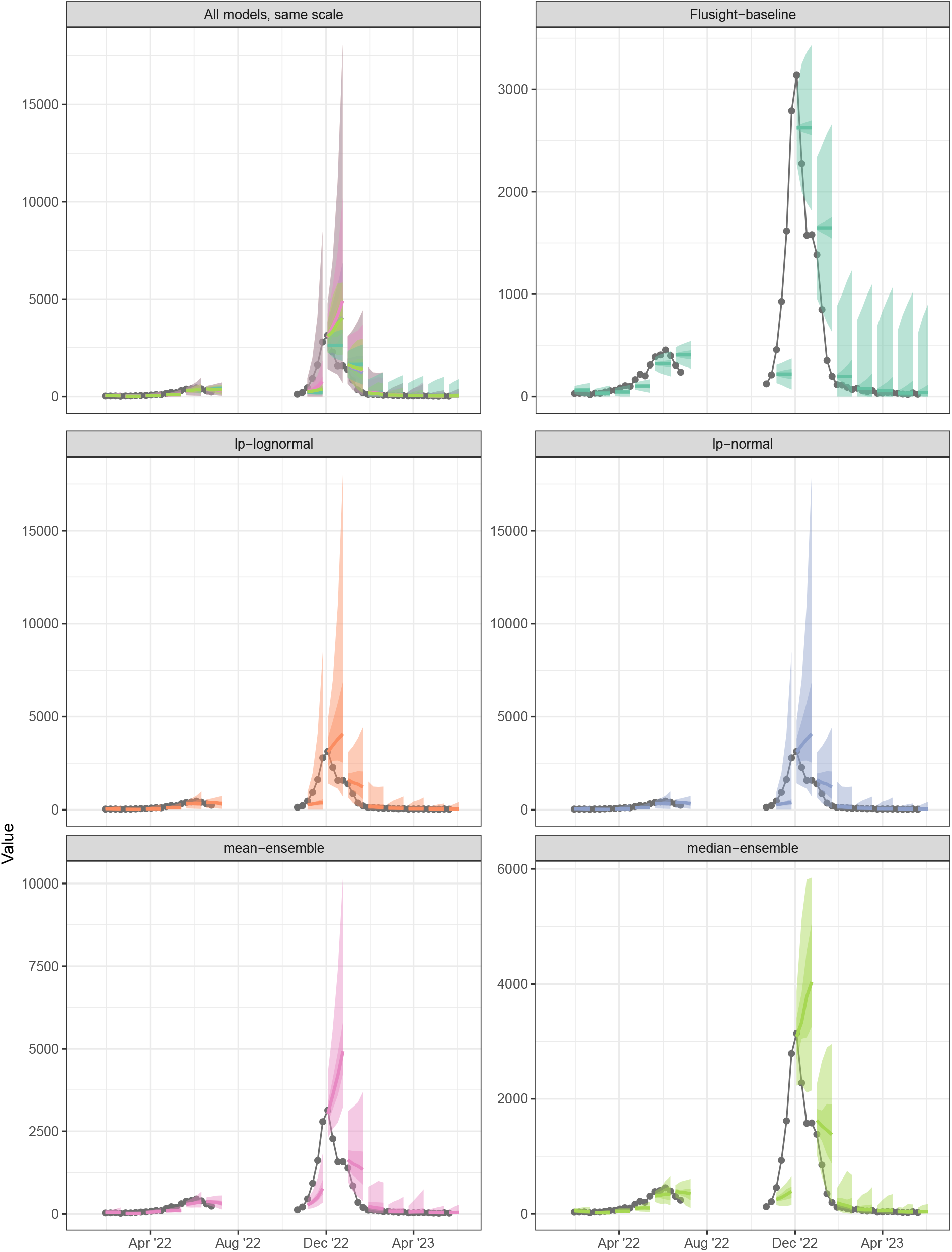
One to four week ahead forecasts for select dates plotted against target data for California. The first panel shows all models on the same scale. All other panels show forecasts for each individual model, with varying y-axis scales, and their prediction accuracy as compared to observed influenza hospitalizations.

Averaging across all time points, the median ensemble has the best scores for every metric. (Note that we only show plots of WIS vs forecast date, faceted by season and horizon, due to similar trends being observed with MAE and 95% prediction interval coverage). The quantile median outperforms the mean ensemble by a similar amount for both WIS and MAE, particularly around local times of change (see Figure 8). The median ensemble also has better coverage rates than the mean ensemble in the tails of the distribution (95% intervals) and similar coverage in the center (50% intervals). The median model also outperforms the linear pools for most weeks, with the greatest differences in scores being for WIS and coverage rates (Figure 8). This seems to indicate that the linear pools’ estimates are usually too conservative, with their wide intervals and higher-than-nominal coverage rates being penalized by WIS. However, during the 2022-2023 season there are several localized times when the linear pools showcased better one-week-ahead forecasts than the median ensemble (Figure 8). These localized instances are characterized by similar MAE values for the two methods and poor median ensemble coverage rates. In these instances, the wide intervals from the linear pools were useful in capturing the eventually-observed hospitalizations, usually during times of rapid change.

**Figure 8:**
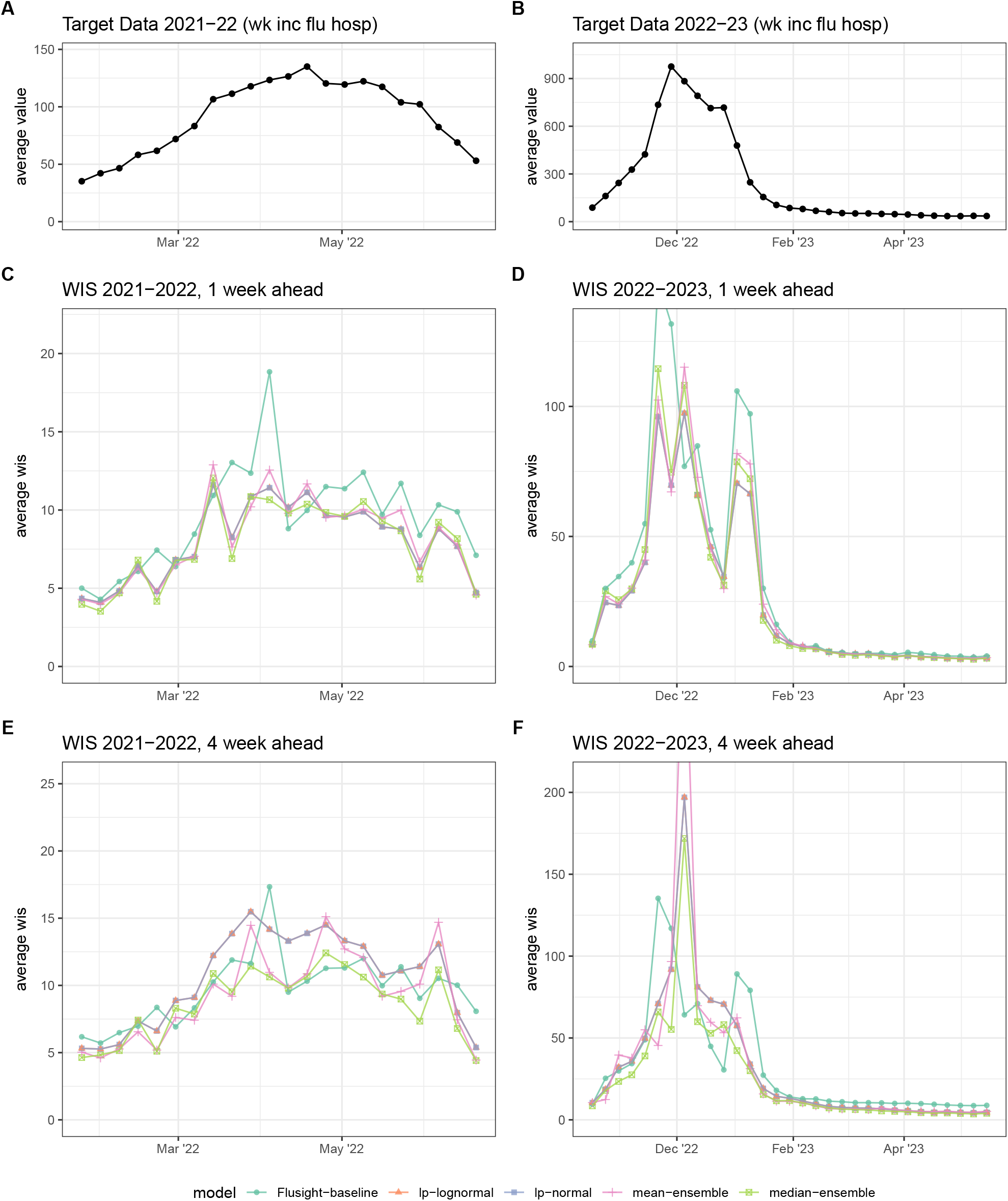
Weighted interval score (WIS) averaged across all locations. Average target data across all locations for 2021-2022 (A) and 2022-2023 (B) seasons for reference. For each season, average WIS is shown for 1-week (C-D) and 4-week ahead (E-F) forecasts. Results are plotted for each ensemble model (colors) across the entire season. Lower values indicate better performance.

All of the ensemble variations outperform the baseline model in this analysis, led by the quantile median which had the best scores overall for WIS, MAE, 50% PI coverage, and 95% PI coverage. However, other models sometimes demonstrated better performance, especially around the season’s peak. These results seem to be consistent with previous findings: linear pools produce intervals generally too wide for a short-term forecasting, except under conditions with greater levels of uncertainty or where the individual models contributing to the ensemble are poorly calibrated. Thus, while the quantile median offers consistency and robustness, there may be certain influenza seasons in which other ensemble methods display better performance.

The choice of an appropriate ensemble aggregation method may depend on the forecast target, the goal of forecasting, and the behavior of the individual models contributing to an ensemble. One case may call for prioritizing high coverage rates while another may prioritize accurate point forecasts. The `simple_ensemble()` and `linear_pool()` functions and the ability to specify component model weights and an aggregation function for `simple_ensemble()` allow users to implement a variety of ensemble methods.

## 6 Summary and discussion

Ensembles of independent models are a powerful tool to generate more accurate and more reliable predictions of future outcomes than a single model alone. Here, we have provided an overview of multi-model ensemble methodology with the goal of improving use of ensemble results in public health and biomedical research settings, as well as other domains that use probabilistic forecasting. Moreover, we have demonstrated how to utilize `hubEnsembles`, a simple and flexible framework to combine individual model predictions into an ensemble.

Multi-model ensembles are becoming the gold standard for prediction exercises in the public health domain. Collaborative modeling hubs can serve as a centralized entity to guide and elicit predictions from multiple independent models, as well as to generate and communicate ensemble results^12,36^. Given the increasing popularity of multi-model ensembles and collaborative hubs, there is a clear need for generalized data standards and software infrastructure to support these hubs. By addressing this need, the hubverse suite of tools can reduce duplicative efforts across existing hubs, support other communities engaged in collaborative efforts, and enable the adoption of multi-model approaches in new domains.

When generating and interpreting an ensemble prediction, it is important to understand the methods underlying the ensemble, as methodological choices can have meaningful effects on the resulting ensemble and its performance. Although there may not be a universal “best” method, matching the properties of a given ensemble method with the features of the component models will likely yield best results^24^. Our case study on seasonal influenza forecasts in the US demonstrates this point. The quantile median ensemble performs best overall for a range of metrics, including weighted interval score, mean absolute error, and prediction interval coverage. Yet, the other ensemble methods we tested also showcased a clear improvement over the baseline model and even outperformed the quantile median for certain weeks, particularly during periods of rapid change when outlying component forecasts are likely more important. The accuracy gains from ensemble models motivate the use of a “hub-based” approach to prediction for infectious diseases, public health, and in other fields.

## Data Availability

All code necessary to reproduce this manuscript can be found in a GitHub repository available at https://github.com/hubverse-org/hubEnsemblesManuscript, including required package versions which are available in the DESCRIPTION file.

https://github.com/hubverse-org/hubEnsembles

## Acknowledgements

The authors thank all members of the hubverse community; the broader hubverse software infrastructure made this package possible. L. Shandross, A. Krystalli, N. G. Reich, and E. L. Ray were supported by the National Institutes of General Medical Sciences (R35GM119582) and the US Centers for Disease Control and Prevention (U01IP001122 and NU38FT000008). E. Howerton was supported by NSF RAPID awards DEB-2126278 and DEB-2220903, as well as the Eberly College of Science Barbara McClintock Science Achievement Graduate Scholarship in Biology at the Pennsylvania State University. L. Contamin and H. Hochheiser were supported by NIGMS grants U24GM132013 and R24GM153920. The content is solely the responsibility of the authors and does not necessarily represent the official views of NIGMS, the National Institutes of Health, or CDC.

## Consortium of Infectious Disease Modeling Hubs

Consortium of Infectious Disease Modeling Hubs authors include Alvaro J. Castro Rivadeneira (University of Massachusetts Amherst), Lucie Contamin (University of Pittsburgh), Sebastian Funk (London School of Hygiene & Tropical Medicine), Aaron Gerding (University of Massachusetts Amherst), Hugo Gruson (data.org), Harry Hochheiser (University of Pittsburgh), Emily Howerton (The Pennsylvania State University), Melissa Kerr (University of Massachusetts Amherst), Anna Krystalli (R-RSE SMPC), Sara L. Loo (Johns Hopkins University), Evan L. Ray (University of Massachusetts Amherst), Nicholas G. Reich (University of Massachusetts Amherst), Koji Sato (Johns Hopkins University), Li Shandross (University of Massachusetts Amherst), Katharine Sherratt (London School of Hygene and Tropical Medicine), Shaun Truelove (Johns Hopkins University), Martha Zorn (University of Massachusetts Amherst)

## Footnotes

1 https://hubverse-org.github.io/hubUtils/reference/as_model_out_tbl.html

2 https://reichlab.io/covidHubUtils/reference/score_forecasts.html

3 https://reichlab.io/covidHubUtils/reference/score_forecasts.html

## Notes

### Competing Interest Statement

The authors have declared no competing interest.

### Summary of Updates

The manuscript has been revised to serve as a tutorial piece, rather than a software manuscript. This includes adding an overview figure (new Figure 1) and a new section (2.3) that describes the application of multi-model ensembles in public health. The other major components of the manuscript remain largely unchanged, with edits for flow and consistency.

